# COVID-19 inequalities in England: a mathematical modelling study of transmission risk and clinical vulnerability by socioeconomic status

**DOI:** 10.1101/2024.01.11.24301159

**Authors:** Lucy Goodfellow, Edwin van Leeuwen, Rosalind M Eggo

## Abstract

**Background:** The COVID-19 pandemic resulted in major inequalities in infection burden between areas of varying socioeconomic deprivation in many countries, including England. Areas of higher deprivation tend to have a different population structure - generally younger - which can increase viral transmission due to higher contact rates in school-going children and working-age adults. Higher deprivation is also associated with higher presence of chronic comorbidities, which were convincingly demonstrated to be risk factors for severe COVID-19 disease. These two major factors need to be combined to better understand and quantify their relative importance in the observed COVID-19 inequalities.

**Methods:** We used UK Census data on health status and demography stratified by decile of the Index of Multiple Deprivation (IMD), which is a measure of socioeconomic deprivation. We calculated epidemiological impact using an age-stratified COVID-19 transmission model, which incorporated different contact patterns and clinical health profile by decile. To separate the contribution of each factor, we also considered a scenario where the clinical health profile of all deciles was at the level of the least deprived. We also considered the effectiveness of school closures in each decile.

**Results:** In the modelled epidemics in urban areas, the most deprived decile experienced 9% more infections, 13% more clinical cases, and a 97% larger peak clinical size than the least deprived; we found similar inequalities in rural areas. 21% of clinical cases and 16% of deaths in England observed under the model assumptions would not occur if all deciles experienced the clinical health profile of the least deprived decile. We found that more deaths were prevented in more affluent areas during school closures.

**Conclusions:** This study demonstrates that both clinical and demographic factors synergise to generate health inequalities in COVID-19, that improving the clinical health profile of populations would increase health equity, and that some interventions can increase health inequalities.

## Background

The COVID-19 pandemic disproportionately affected people in lower socioeconomic groups around the world [1], [2]. In England, there were large disparities in COVID-19 burden between areas of different relative deprivation, measured by the Index of Multiple Deprivation (IMD). Initial reports by the UK Office for National Statistics (ONS) found that from April-July 2020 the most deprived 10% of areas in England experienced an age-standardised COVID-19-related mortality rate more than twice as high as the least deprived 10% [3]. These disparities were repeatedly observed through the pandemic: between June 2020 and January 2021, the age-standardised mortality rate in laboratory-confirmed cases of COVID-19 was 371.0 per 100,000 (95% Confidence Interval (CI): 334.2 - 410.7) compared to 118.0 (95% CI: 97.7-141.3) in the most vs least deprived quintiles [4]. The inequality in mortality rates seen in the early pandemic exceeded that observed in previous years, indicating that there were further factors exacerbating the ‘expected’ effects of relative deprivation [5]. Even after adjusting for age, sex, region, and ethnicity, this report found worse outcomes in more deprived areas, but did not adjust for prevalence of comorbidities. Other studies have consistently confirmed an association between comorbidities and more severe COVID-19 outcomes [6], [7].

Morbidity and the presence of underlying health conditions tend to vary greatly by socioeconomic status (SES), and are a significant risk factor for severe infection [8]. Vulnerability to more severe infection has both direct effects, including a greater risk of consequential long-term health complications and greater mortality risk, and indirect population-level effects, such as potentially increased infectiousness of symptomatic cases. In England, before the COVID-19 pandemic, life expectancy was 9.4 years longer for men in the least deprived decile than the most, and 7.7 years longer for women [9]. These gaps continue to widen: female life expectancy in the most deprived decile fell by 4 weeks between 2014-2016 and 2017-2019, but rose by 11 weeks in the least deprived [9], [10]. The prevalence of underlying health conditions that affect quality of life is also consistently correlated with local deprivation levels: men in the most deprived decile could expect to live 18.4 years fewer in good health than those in the least deprived decile; the corresponding gap for women was 19.8 years [9].

It is well established that infectious disease burden is associated with SES [11]–[13]. This is linked to a multitude of complex and interwoven factors including, but not limited to, lack of access to healthcare, poor housing conditions, inability to avoid high-exposure settings such as crowded public places, differences in occupation type, and avoiding restrictions or testing due to mistrust of authorities [14], [15].

Here, we use a novel transmission model to combine differences between socioeconomic groups in their risk of infection with their risk of severe disease on infection to quantify their relative importance in the observed COVID-19 inequalities in England. We consider underlying health conditions as a key determinant of an individual’s risk of developing a clinical case of COVID-19, and focus on the impact of IMD-specific health and age structure on infectious disease burden at the population level. By making simplifying assumptions and modelling a synthetic population, we aim to produce a conceptual exploration of the interaction between underlying health and demographic structure.

## Methods

We developed an age-stratified dynamic transmission model for SARS-CoV-2, which was further stratified by IMD decile, and by urban or rural classification in England. Here we detail how the model was modified to incorporate the characteristics of each decile and geography.

## IMD-specific age structure

Each epidemic was simulated on the population of a given IMD decile in either an urban or rural area, to account for the distinct underlying age structures in these areas. We used 17 age groups (0-1, 1-5, every 5 years to 75, and over 75). The mid-2020 (30 June) age-specific population of each lower layer super output area (LSOA), which is on average 1,500 people, was linked via LSOA codes to their IMD decile and urban/rural classification (where urban is defined as a settlement with over 10,000 residents) [16]–[19]. We calculated the size of each age group, specific to each IMD decile and geography, and used this to determine the average age structure of each IMD- and geography-specific population, *n* = (*n*_1_, …, *n*_17_) where 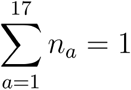 in each population. We also calculated the median age for each urban and rural IMD decile, and the proportion of each IMD decile residing in urban or rural LSOAs (Supplementary Section 1).

### Contact matrices

To define contact between age groups we used age-specific social contact data for the United Kingdom (UK) for physical and conversational contacts accessed via the *socialmixr* R package [20], [21]. The contact matrices are highly age-assortative, with the highest daily contact patterns occurring between individuals in the same age group for those aged 5-19. We projected the contact patterns onto the age structure of each population in 2020, using the density correction method, by constructing an intrinsic connectivity matrix and scaling this matrix to match the population’s age structure [22].

The intrinsic connectivity matrix was calculated from the 2006 UK contact matrix 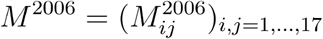 and age structure 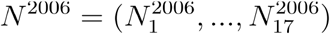 as

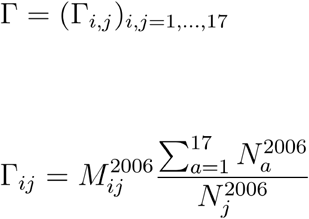

The new contact matrix for a population with age group sizes *N* = (*N*_1_, …, *N*_17_) and proportions *n* = (*n*_1_, …, *n*_17_) had entries

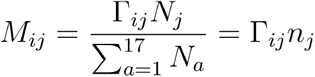

### Age-specific fraction of COVID-19 cases causing clinical symptoms

We separate infections of SARS-CoV-2 as in [23], into clinical or subclinical cases. Clinical cases of COVID-19 are infections that lead to noticeable symptoms such that an individual may seek clinical care. Subclinical infections do not seek care in the model, and are less infectious than clinical cases. We defined a population’s clinical fraction as the probability of an individual in the population developing a clinical case of COVID-19 upon infection. Here, we extended previous work by relating an individual’s probability of being a clinical case of COVID-19 to the self-reported health status of their IMD- and age-specific population in England, and then examined how differences in self-reported health status by IMD decile, coupled with differences in age distribution, affect the burden in each IMD decile.

To define health status, we used data from the 2021 Census, specifically the question ‘How is your health in general?’, with response options of ‘Very good’, ‘Good’, ‘Fair’, ‘Bad’, and ‘Very bad’ [24]. This is provided by the Census stratified by IMD and by age. We then defined ‘health prevalence’ as the proportion of individuals reporting ‘Very good’ or ‘Good’ general health, stratified by the same age groups and the deciles of IMD:

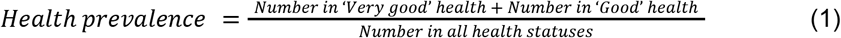

To map the population health prevalence to the model’s age-specific clinical fraction we used locally weighted least squares regression (LOESS) [23]. Any populations with health prevalences outside of the training dataset’s range were assigned the most extreme clinical fractions found by Davies et al. [23], to avoid extrapolation outside of observed values. Health prevalence was highest in children, but children have separate risk factors for severe disease (such as smaller airways) and children under 10 have been found to be subject to a higher risk of clinical COVID-19 cases and a greater infection fatality ratio (IFR) [23], [25] (as observed for other infections such as influenza [26]). Therefore, we fixed the clinical fraction of the 0-9 age group at 0.29, matching that found by [23].

### COVID-19 transmission model

The transmission model includes a single SARS-CoV-2 variant, no existing immunity in the population, and natural history parameters drawn from the first wave of the pandemic. We did not include vaccination, and the only non-pharmaceutical intervention (NPI) considered was school closures. We developed an age- and IMD-stratified deterministic compartmental model in R (version 4.3.1) (Figure 1c). There is no mixing between IMD deciles in the model. The aim is to demonstrate the importance of health prevalence and differences in age and social mixing in epidemic impact, rather than to reproduce the COVID-19 epidemic in England.

**Figure 1.**
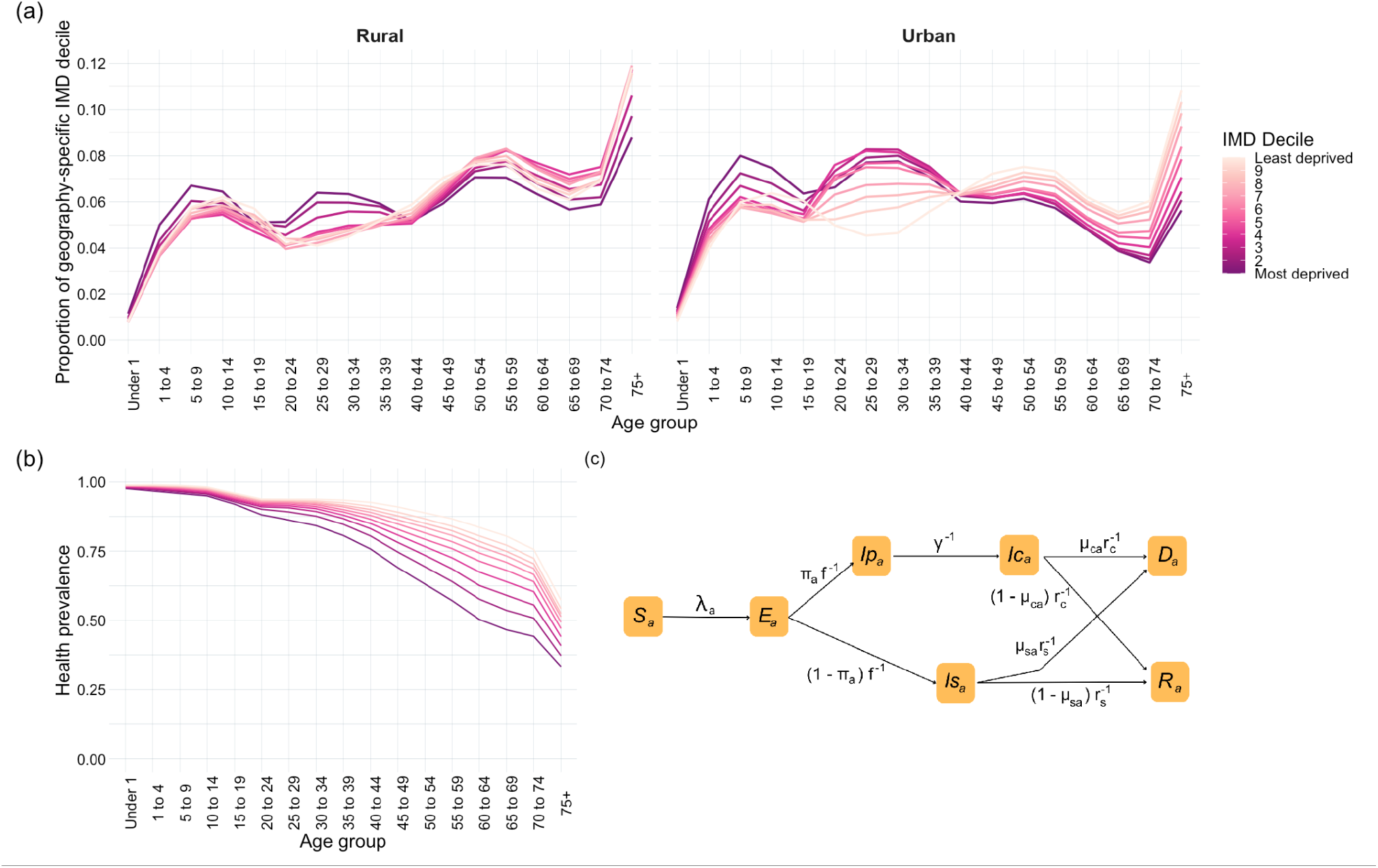
**a)** Proportion of each geography-specific IMD decile in each age group. **b)** Age- and IMD-specific health prevalence (1 most deprived decile, 10 least deprived). **c)** Age-stratified SEIRD model, specific to IMD decile and geography. Subscript *a* denotes age-specificity, *c* clinical parameters, and *s* subclinical parameters.

Individuals are first assumed to be susceptible (*S*), and become exposed (*E*) but not yet infectious after effective contact with an infected individual (Figure 1c). Each exposed individual then progresses to one of two infected states: subclinical infection (*Is*), and clinical infection, which is represented by a pre-symptomatic (but infectious) compartment (*Ip*) followed by a symptomatic compartment (*Ic*). Each individual then moves into the recovered (*R*) or dead (*D*) compartment, at which point they are assumed to no longer be infectious and to be immune to infection. This Susceptible-Exposed-Infectious-Recovered-Dead (SEIRD) is an extension of [23], with the addition of a *D* compartment. We ran the epidemic for 365 days, which allowed completion of each epidemic in each decile and geography. Each epidemic was run on a synthetic population of a fixed IMD decile and urban/rural geography, with no births, non-infection-related deaths, or ageing between age groups, as the time-frame of each epidemic was less than a year. The model also assumed that contact patterns remain constant through the epidemic.

The force of infection in age group k is given by:

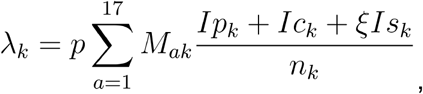

where *p* is the probability of a contact between an infected and susceptible individual resulting in transmission of infection, *M* _*ak*_ is the mean daily number of contacts that an individual in age group *a* has with individuals in age group k, and ξ is the relative infectiousness of subclinical cases. The age-specific clinical fraction is denoted by π_*a*_ and depends on the IMD decile. Rates of transition from each disease state are given in Table 1.

**Table 1.** Model parameters.

| Parameter | Value | Definition | Source |
| --- | --- | --- | --- |
| $p$ | 0.06 | Transmission probability | [23] |
| $M_{ak}$ | Varies by age and IMD | Daily age-specific contacts | Based on [20] |
| $\xi$ | 0.5 | Relative subclinical infectiousness | Assumption |
| $\pi_a$ | Varies by age and IMD | Clinical fraction | Based on [23] |
| $f$ | 3 days | Duration of latent period | [23] |
| $\gamma$ | 2.1 days | Duration of preclinical infectious period | [23] |
| $r_c$ | 2.9 days | Duration of clinical infectious period | [23] |
| $r_s$ | 5 days | Duration of subclinical infectious period | [23] |
| $\mu_{ca}$ | Varies by age, see Supplementary Table 4 | Clinical mortality probability | [27] |
| $\mu_{sa}$ | 0 for all age groups | Subclinical mortality probability | Assumption |

We assumed the relative subclinical infectiousness (ξ), to be equal to 0.5, and tested this assumption in a sensitivity analysis (see Supplementary Section 12). The transmission probability during a contact was assumed to be *p* = 0. 06 as in [23]. Remaining parameter estimates were taken from [23] where possible, to replicate the conditions used to derive the clinical fraction estimates. The mortality probability of subclinical infections was assumed to be 0 for all age groups (*a*). The age-specific probability of mortality of clinical cases were estimated using age-specific IFRs (*ϕ* _*a*_) found by Verity et al. in 2020 [27] (Supplementary Table 4). As the IFR is, *ϕ*_*a*_ *= π*_*a*_ *μ*_*ca*_ + (1 − *π*_*a*_) *μ*_*sa*_ *=* π_*a*_μ_*ca*_, since *μ*_*sa*_ the age-specific clinical mortality probabilities were estimated by

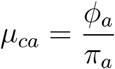

where π_*a*_ are the age-specific clinical fractions for the general population in [23] (Supplementary Table 4).

We calculated the total infections, clinical cases, and fatalities per 1,000 people, the peak number of clinical cases per 1,000 people, the IFR, and the basic reproduction number (*R*_*0*_) for each IMD decile in urban and rural areas. We also calculated age-standardised measures of total infections, clinical cases, and fatalities within a specific geography for increased comparability. The age-standardised results were of the form:

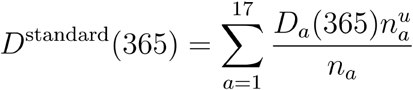

Where 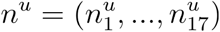 is the standard urban population, similarly 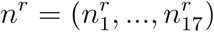 for rural areas.

*R*_*0*_ of each IMD decile in urban and rural areas was calculated as the absolute value of the largest eigenvalue of the next-generation matrix *N*:

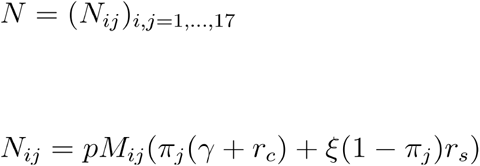

### Counterfactual scenarios

To determine the epidemic burden attributable to the difference in underlying health status between IMD deciles, we created the counterfactual health prevalence scenario, where all deciles were assigned the age-specific health prevalence of decile 10 (the least deprived). We calculated the total clinical cases and fatalities in each IMD decile under this assumption. In order to reflect the size of each population (while each IMD decile comprises 10% of the population of England, geography-specific IMD deciles vary widely in size, see Supplementary Table 1), we scaled mortality to mid-year 2020 population sizes and totalled over the 20 populations.

We also created the counterfactual scenario of constant age structure, where we held the age structure constant at the average of each geography-specific England population, independent of IMD decile. This allowed us to determine the impact of clinical vulnerability separately from the differences in age distribution in each IMD decile. The health prevalence by age remained at the IMD-specific value.

### School closures

To quantify potential differences in impact of school closures in different IMD deciles, we calculated the effect of school closures on *R*_*0*_ and total fatalities. We removed the school-specific contacts from the contact matrix (retaining contacts in home, work, leisure, transport, or other locations), re-projected onto the 2020 age structure, and recalculated the next-generation matrix, *N*, and its largest eigenvalue, *R*_*0*_. While assuming that closure of schools results in a complete subtraction of school-specific contacts may not be realistic (as some contacts would likely be replaced by social interactions in other locations [28]), the results demonstrate the maximum potential impact of school closures.

We simulated closure of schools after a certain cumulative proportion, P, of the population developed clinical COVID-19 cases. The use of cumulative clinical cases as a threshold for implementation is reflective of using total confirmed cases as a measure of the size of an early epidemic. We assumed a value of P = 0.05, but tested different values in sensitivity analyses (Supplementary Section 11).

## Results

### Self-reported health prevalence is lower in more deprived areas

There was an older age structure in rural areas compared to urban, and a generally younger age structure in more deprived areas (Figure 1a). The relationship between IMD decile and age structure was confirmed by the median age in each population (Supplementary Figure 1); rural areas have consistently higher median ages than urban areas of the same IMD decile. Median age monotonously increased with affluence in urban areas, but peaked in the fourth decile for those living in rural areas.

Health prevalence was lower in each IMD decile as age increased, and lower in every age group as relative deprivation increased (Figure 1b). 47% of those aged 65-69 living in the most deprived decile reported living in ‘Very good’ or ‘Good’ health, compared to 80% of those in the least deprived decile. Those living in the most deprived decile experienced the same health prevalence (76%) at age 40-44 as those in the least deprived decile did at age 70-74.

Health prevalence was mapped to clinical fraction in the age groups used in [23] as described in Methods (Figure 2a). Under this assumption, all those over the age of 10 in more deprived areas had a greater likelihood of developing a clinical case of COVID-19 than in other deciles (Figure 2b).

**Figure 2.**
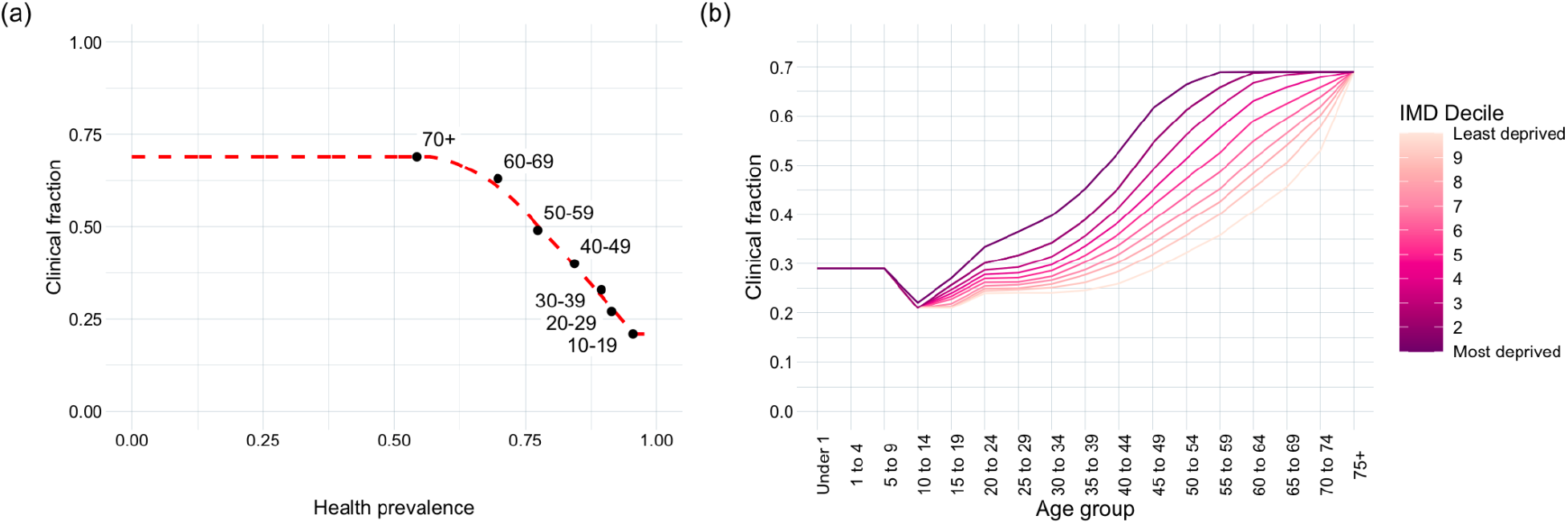
Results of mapping underlying health to clinical vulnerability. **a)** The training dataset of age-specific health prevalence and clinical fraction estimates for the general population of England over age 10, and corresponding LOESS smoother, with linear extensions outside the domain [0.21, 0.69]. **b)** Resulting age- and IMD-specific clinical fractions (1 most deprived decile, 10 least deprived).

### Epidemic burden increases with relative deprivation

We found that total infections and clinical cases increased with deprivation (Figure 3a, b). In rural settings, the most deprived decile experienced 72 more crude infections per 1,000 population than the least deprived decile; this inequality increased to 90 infections in urban settings. The inequalities in clinical cases were even larger: in rural areas, the most deprived decile experienced 147 more clinical cases per 1,000 than the least, and 130 more clinical cases in urban areas. The peak clinical epidemic size was 97% larger in urban areas of the most deprived decile than the least deprived decile under these model assumptions, and 91% larger in rural areas (Figure 3c).

**Figure 3.**
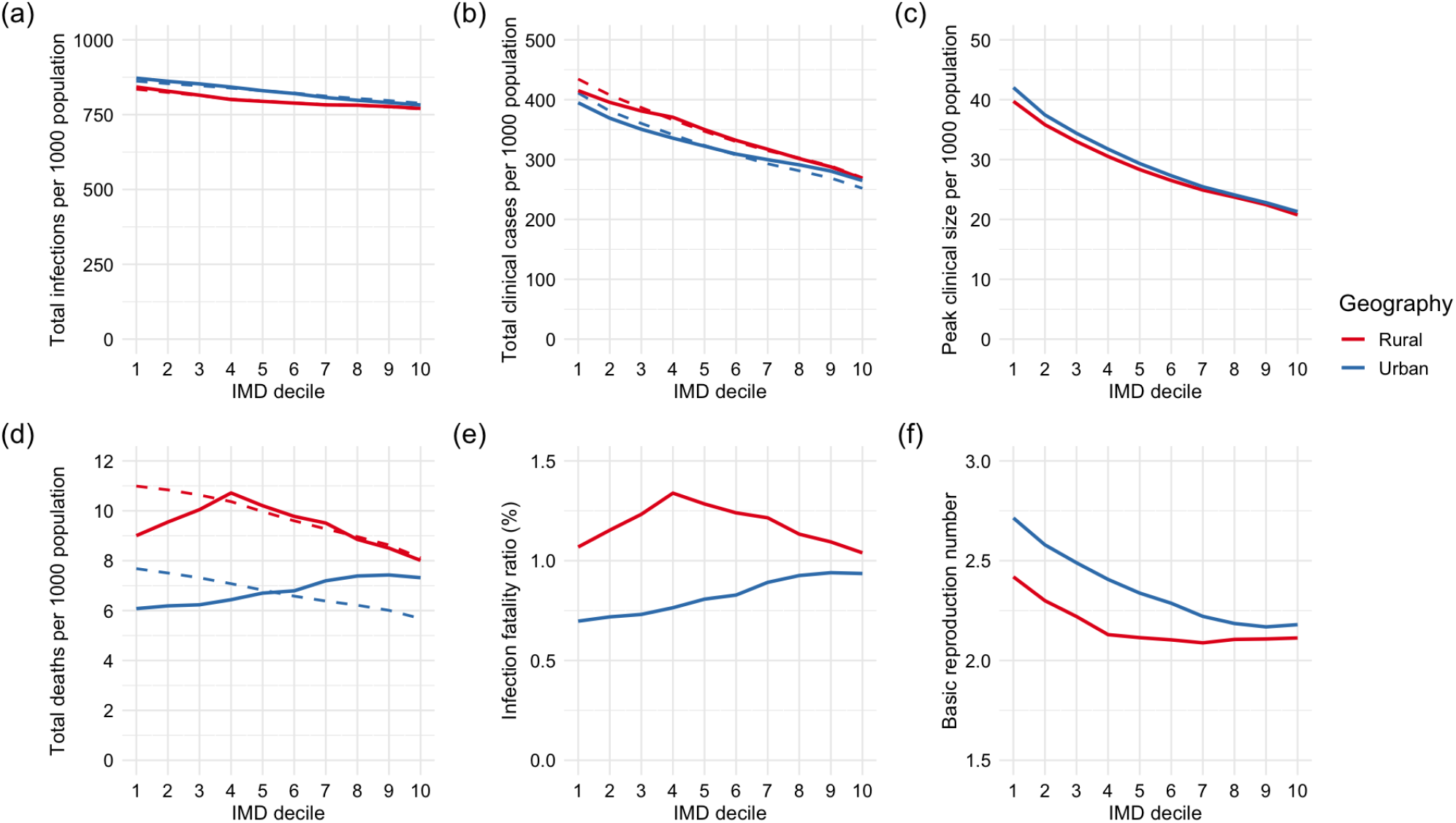
Measures of size of a COVID-19 epidemic in each IMD decile and geography. Solid lines represent crude measures, and dashed lines those age-standardised by geography. The most deprived decile is decile 1, and the least is decile 10. **a)** Total infections per 1,000 population. **b)** Total clinical cases per 1,000 population. **c)** Clinical cases per 1,000 population at the clinical peak of the epidemic. **d)** Total deaths per 1,000 population. **e)** Infection fatality ratio. **f)** Basic reproduction number, *R*_*0*_.

Mortality inequalities differed between the crude and age-standardised results (Figure 3d). The crude total number of deaths by IMD decile and geography closely followed the median age (Supplementary Figure 1). There was a strong positive association between increasing relative deprivation (decreasing decile) and the age-standardised number of deaths (Figure 3d). In urban areas, 2.0 more deaths occurred per 1,000 age-standardised population in the most deprived decile than the least; this inequality increased to 2.9 deaths per 1,000 age-standardised population in rural areas. The IFR followed a very similar pattern to crude mortality (Figure 3e), likely due to a combination of the relative stability of total infections with deprivation compared to the large variation in mortality rates, and the strong relationship between median age and mortality.

*R*_*0*_ was generally higher in more deprived areas (Figure 3f), and ranged from 2.09 in rural areas of the 7th decile to 2.71 in urban areas of the most deprived decile. The *R*_*0*_ was not strongly related to median age because the lower clinical fractions in younger populations was counteracted by their higher contact rates.

Rural areas experienced fewer total infections, lower peak clinical sizes, and lower *R*_*0*_ than urban areas, but more clinical cases and deaths, at all levels of deprivation. This is likely due to the older rural age structure, as older individuals had fewer daily contacts than younger individuals and so produced fewer secondary infections, but were more likely to develop clinical COVID-19 if infected.

### Health-attributable deaths occur at all ages

Under the counterfactual health prevalence scenario, 340,532 deaths occurred, compared to the 405,695 under the original assumption. Therefore, 16% of deaths, or over 65,000 fatalities, would have been prevented by achieving health prevalence equity at the level of the least deprived decile. These health-attributable deaths did not only occur in those at older ages: over 29,000 prevented deaths were in individuals aged under 65 (Supplementary Figure 6). At all ages between 30 and 70, over 20% of deaths that occurred under the original model assumptions were attributable to underlying health inequalities (Supplementary Figure 7). We similarly found 21% of clinical cases (3.8 million) to be attributable to inequalities in underlying health under the model assumptions.

Lower clinical infection and mortality rates occurred in the most deprived areas, in both urban and rural geographies in the counterfactual health prevalence scenario (Figure 4). We also found that age-standardised deaths were consistent across IMD deciles in both geographies under the counterfactual health prevalence scenario (Supplementary Figure 10). This result contradicts observed mortality rates [3], [4], providing evidence for the existence of a dependency of clinical vulnerability on IMD and more specifically underlying health. The true relationship between IMD and age-specific clinical fraction may be more complex than the assumptions made in this paper; for example, pre-existing immunity may be dependent on previous exposure to coronaviruses [29], which may be associated with SES but is not considered here.

**Figure 4.**
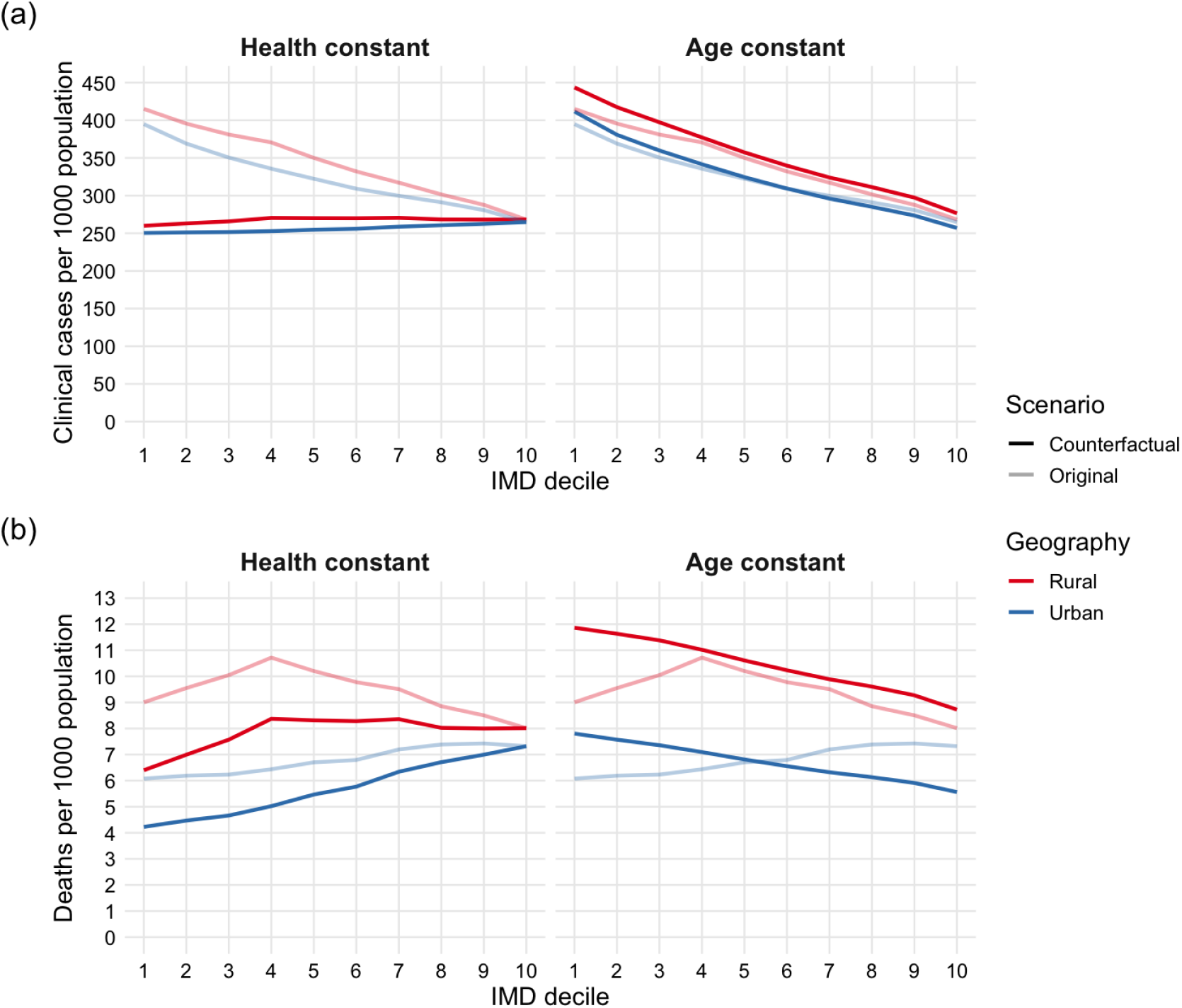
Epidemiological burden in counterfactual scenarios. **a)** Total clinical cases per 1,000 population, in geography-specific areas of each IMD decile (1 most deprived decile, 10 least deprived), in the counterfactual health prevalence scenario, and the counterfactual constant age structure scenario. The original model is shown for comparison in pale lines. **b)** Total deaths per 1,000 population, in geography-specific areas of each IMD decile, under the same scenarios.

In the counterfactual scenario of constant age structure, we observed more clinical cases and deaths in more deprived areas (Figure 4). We also considered an underlying age structure independent of IMD decile or geography and found that the most deprived decile experiences 40% higher mortality and a clinical peak 1.88 times larger than the least deprived decile (Supplementary Figure 5), demonstrating the inequality resulting from health prevalence separately from demographic differences. These results indicate that observed inequalities in clinical case numbers and mortality are the result of a complex interaction between comorbidity-related clinical vulnerability and a population’s demographic structure, the outcome of which is not necessarily consistently related to deprivation.

Further sensitivity analyses considering epidemiological parameters show consistent patterns of age-standardised mortality by deprivation, but a change in the pattern of crude deaths as epidemiological parameters are varied (Supplementary Section 12).

### School closures prevent more deaths in less deprived areas

With school closures in place in the model, *R*_*0*_ decreased for all geographies and IMD deciles, but remained larger in urban than rural areas and was consistently higher in more deprived areas in both geographies (Figure 5a). In urban areas, *R*_*0*_ was 0.38 higher in the most deprived decile than the least; the equivalent inequality was 0.29 in rural areas. The largest reductions in *R*_*0*_ occurred in the most and least deprived deciles, with the least impact in the median deciles (Figure 5b). This U-shaped result is likely a product of the age structure of each population, as *R*_*0*_ is driven by both high daily contact patterns in young individuals and greater clinical vulnerability in older individuals (more detail in Supplementary Section 11). In all IMD deciles, greater reductions in *R*_*0*_ occurred in urban than rural areas, likely due to the greater proportion of school-aged children and hence larger reduction in contacts. In no scenario was *R*_*0*_ reduced below 1 (Figure 5a), meaning that school closures were not able to halt COVID-19 transmission in any rural or urban IMD decile and could only reduce epidemic burden under our model assumptions.

**Figure 5.**
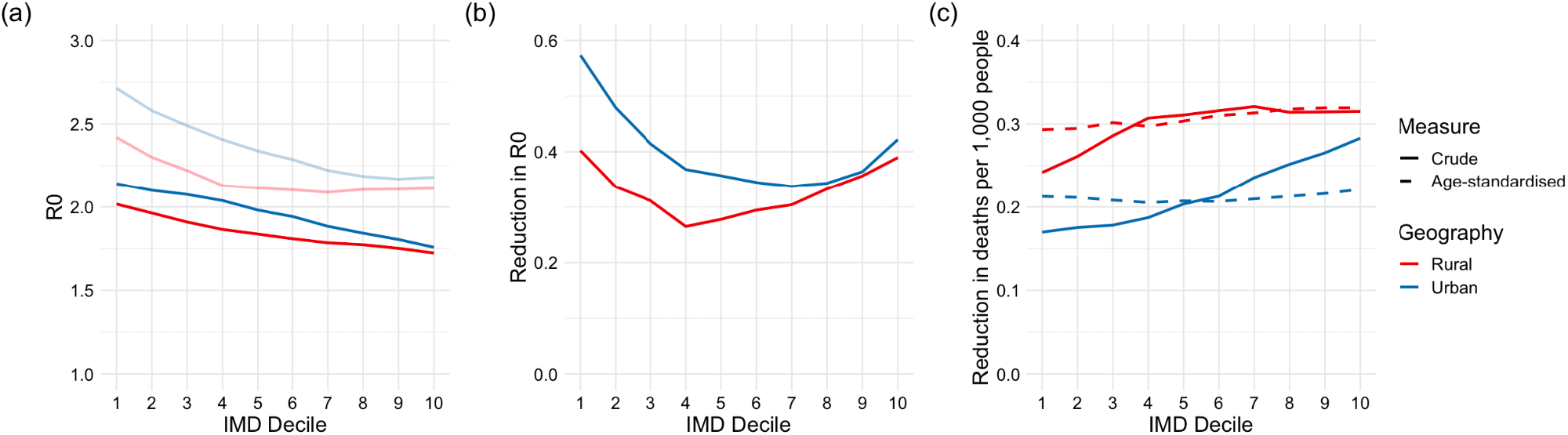
Results of implementing school closures. **a)** *R*_*0*_ in each IMD- and geography-specific population (1 most deprived decile, 10 least deprived), before (pale lines) and after school closures. **b)** Reductions in *R*_*0*_ due to school closures. **c)** Crude (solid lines) and age-standardised by geography (dashed lines) reductions in deaths observed per 1,000 population after implementing school closures at P = 0.05.

By implementing school closures after 5% of the population experienced a clinical case of COVID-19 (P = 0.05), 0.113 more crude deaths were prevented per 1,000 people in the least deprived urban areas than the most deprived, with a corresponding difference of 0.073 deaths per 1,000 people in rural areas (Figure 5c). This is likely due to a combination of more crude deaths occurring in more affluent deciles without intervention, improved health conditions, and older population structures. The deaths prevented when age-standardised by geography, shown as dashed lines, were approximately consistent by IMD. We also investigated the pattern of prevented mortality when changing the school closure implementation threshold (Supplementary Section 11) and found that the effectiveness of school closures in less deprived areas decreased dramatically as P increased.

## Discussion

We have shown that, under the assumption that vulnerability to clinical COVID-19 infection is a direct result of a population’s health prevalence, total COVID-19 infections, clinical cases, and age-standardised deaths consistently increased with relative deprivation, therefore exposing those living in the most deprived areas to a greater risk of mortality, as well as more non-fatal consequences such as hospitalisation and long COVID. The peak clinical size of the modelled COVID-19 epidemics, which describes the worst-case scenario hospitals would have to withstand, was approximately twice as large in the most deprived decile than the least deprived. We have found that 16% of the deaths observed under the assumptions of this model, or over 65,000 deaths, would be prevented if every IMD decile experienced the same age-specific health as the most affluent 10% of the country. We have also shown that school closures, which disproportionately negatively affect children’s education and well-being in more deprived areas, may also disproportionately benefit the most affluent in society in terms of epidemiological burden [30], [31].

This study used publicly available data and relied on simplified models of infectious disease transmission; there are hence several limitations to the study. The self-reported nature of the census data means that there may be systematic differences in how health is reported between age groups and levels of deprivation, due to social desirability and the acceptability of self-reporting ill-health varying by demographic, cultural and socioeconomic factors [9]. Census data and the IMD may exclude mobile communities and the over 270,000 homeless individuals in England, who are often among the most vulnerable members of society [32], [33].

Self-reported health in 2021 may include effects of the COVID-19 pandemic, and so pre-emptively confirm the inequities that this model aims to investigate. However, the IMD-specific health prevalence in 2021 (Supplementary Table 3) is very similar to that found in the 2011 UK Census (75.0% health prevalence in the most deprived decile, and 86.9% in the least deprived decile) [34].

Much of the data used in calculating the IMD relate to 2015-2016 [16]. Any changes that have occurred since are therefore not accounted for in the IMD rankings, such as the wider roll-out of Universal Credit, which has been shown to have exacerbated existing inequalities and negatively impacted claimants’ well-being [35], [36]. Health is itself a component of the IMD, potentially limiting the IMD as an exposure for studies with health outcomes; a brief analysis confirms that there are associations between domains of deprivation other than health (Supplementary Figure 13). Other studies have also confirmed the relationship between local deprivation and health outcomes when factoring out the health component of IMD [37].

The assumption of a closed population is unrealistic: apart from during the most stringent lockdowns, which are not represented by the contact patterns used in the above work, individuals will interact and transmit infection between LSOAs as well as within them. A major limitation of the contact patterns used is that intrinsic contact patterns are unlikely to be constant across all IMD deciles and urban and rural geographies. Contact patterns also drastically change in an epidemic, to an extent which depends on SES. The more affluent can more readily reduce their mobility and exposures, while many in the most deprived deciles have less control over their mobility and exposure patterns and are more likely to be in public-facing employment [38]. The ability to self-isolate may also depend on SES, for instance through the conditions of sick pay. The assumptions of constant contact patterns were necessary due to a lack of readily available data on IMD- and age-specific contact patterns, both under NPIs and in daily life, and as a consequence this study is likely to have underestimated the socioeconomic inequalities in epidemic burden. SES-specific contact patterns should be incorporated into epidemic models to include the different contacts that for example arise from different occupational prevalences, ability to reduce mobility, household size, and classroom size. To this end, further data should be collected and made accessible for future research.

By restricting clinical fractions between 0.21 and 0.69, clinical fractions converged at the upper bound in deprived deciles over age 60 while health prevalences were still diverging, meaning that the assigned clinical fractions may underestimate the potential difference in vulnerability, and therefore epidemiological burden, between these IMD deciles. The parameters used for the model, taken from [23] and [27], contain some uncertainty which is included in the original papers but not considered in this study.

The presence of drastically worse underlying health conditions in more deprived areas of England has caused, and will continue to cause, dramatic inequities in the burden of infectious disease. This study has quantified the potential inequities in epidemic burden under the assumption that vulnerability to severe infection is a direct result of existing comorbidities. The most effective way to reduce the inequality of epidemic burden caused by socioeconomic health inequalities is to improve socioeconomic equity in health in England. The recommendations made by Health Equity in England: The Marmot Review 10 Years On [10], including maximising empowerment for all, improving standards of living, creating fair employment, and developing healthy communities, would reduce avoidable inequalities in health and by extension avoidable inequalities in epidemic burden.

## Data Availability

All analysis code and data are available online at

https://github.com/1035825/imd-covid

## Abbreviations

CI: Confidence Interval
IFR: Infection Fatality Ratio
IMD: Index of Multiple Deprivation
LSOA: Lower Layer Super Output Area
NPI: Non-Pharmaceutical Intervention
SEIRD: Susceptible-Exposed-Infectious-Recovered-Dead
SES: Socio-Economic Status
UK: United Kingdom

## Declarations

### Ethics approval and consent to participate

Not applicable.

### Consent for publication

Not applicable.

### Availability of data and materials

All analysis code and data are available at https://github.com/1035825/imd-covid.

### Authors’ contributions

LG and RME conceived the study. LG developed the mathematical model and conducted the analyses. RME and EvL consulted on the analyses. LG and RME wrote the manuscript. All authors read and approved the final manuscript.

## Acknowledgements

Not applicable.

