## Supplementary Material for "COVID-19 inequalities in England: a mathematical modelling study of transmission risk and clinical vulnerability by socioeconomic status"

Code availability:

<https://github.com/1035825/imd-covid>

### Contents:

1. Age structure
2. Contact matrices in IMD deciles 1 and 10
3. Age- and IMD-specific health statuses
4. Overall health prevalence in each IMD decile
5. Training dataset
6. Clinical mortality probabilities
7. Model equations
8. Epidemic trajectories
9. Counterfactual scenario of constant age structure
10. Counterfactual scenario of underlying health equity
11. School closures
12. Epidemiological sensitivity analyses
13. Association between health prevalence and IMD domains at LSOA level

### 1. Age structure

The median age in each geography- and IMD-specific population (Supplementary Figure 1) confirms the relationship between these factors and age structure observed in Figure 1a; rural areas have consistently higher median ages than urban areas of the same IMD decile. Median age monotonously increases with affluence in urban areas, ranging from 35 to 46, but peaks in the fourth decile for those living in rural areas at age 51.

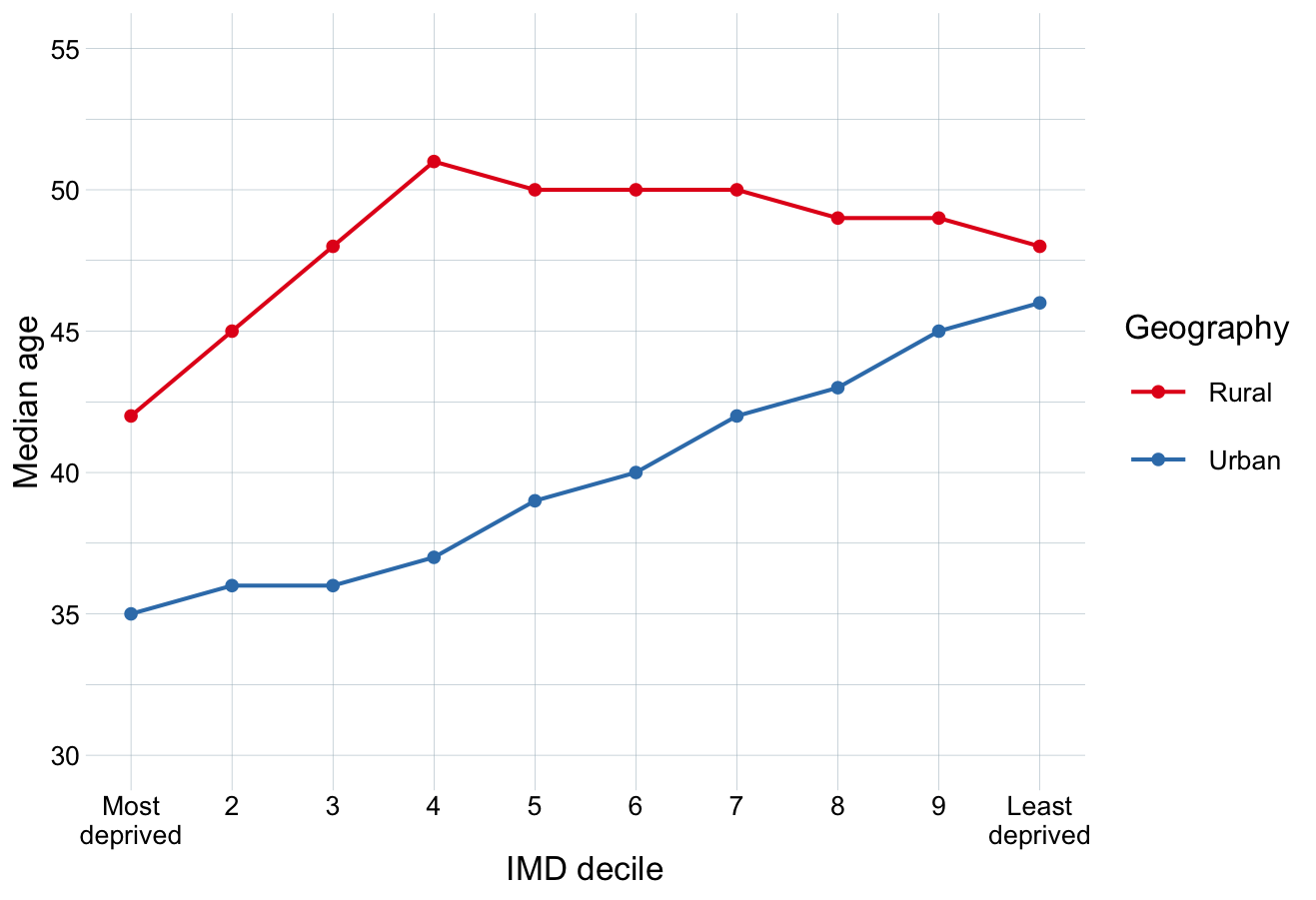

**Supplementary Figure 1.** Median age of each IMD decile, stratified by geography.

While urban areas make up only 15% of England’s land area, they comprise 83% of the population. The proportion of the population in each IMD decile living in urban areas varies greatly: while 98% of those living in the most deprived decile live in urban LSOAs, this falls to 73% for some less deprived deciles (Supplementary Table 1). This means that while 12% of those living in urban areas live in the most deprived decile, less than 1% of those living in rural areas do.

| **IMD decile** | 1 (Most deprived) | 2 | 3 | 4 | 5 | 6 | 7 | 8 | 9 | 10 (Least deprived) |
| --- | --- | --- | --- | --- | --- | --- | --- | --- | --- | --- |
| **Urban proportion** | 0.98 | 0.96 | 0.93 | 0.85 | 0.78 | 0.73 | 0.73 | 0.75 | 0.75 | 0.81 |

**Supplementary Table 1.** Proportion of each IMD decile’s population residing in urban LSOAs.

### 2. Contact matrices in IMD deciles 1 and 10

Supplementary Figure 2 shows the contact matrices produced by projecting the POLYMOD UK contact matrix onto the age structure of each geography-specific IMD decile. Contact patterns are visibly highly age-assortative (on the central diagonal), with the highest daily contact patterns occurring between individuals in the same age group for those aged 5-19. This is accentuated in the most deprived decile, particularly in urban areas, as there is a larger proportion of these age groups present (see Figure 1a, Supplementary Figure 1). There are also increased contact patterns between individuals 25 - 40 years old and those aged 0 - 20 (secondary diagonals), likely representing intergenerational familial contacts.

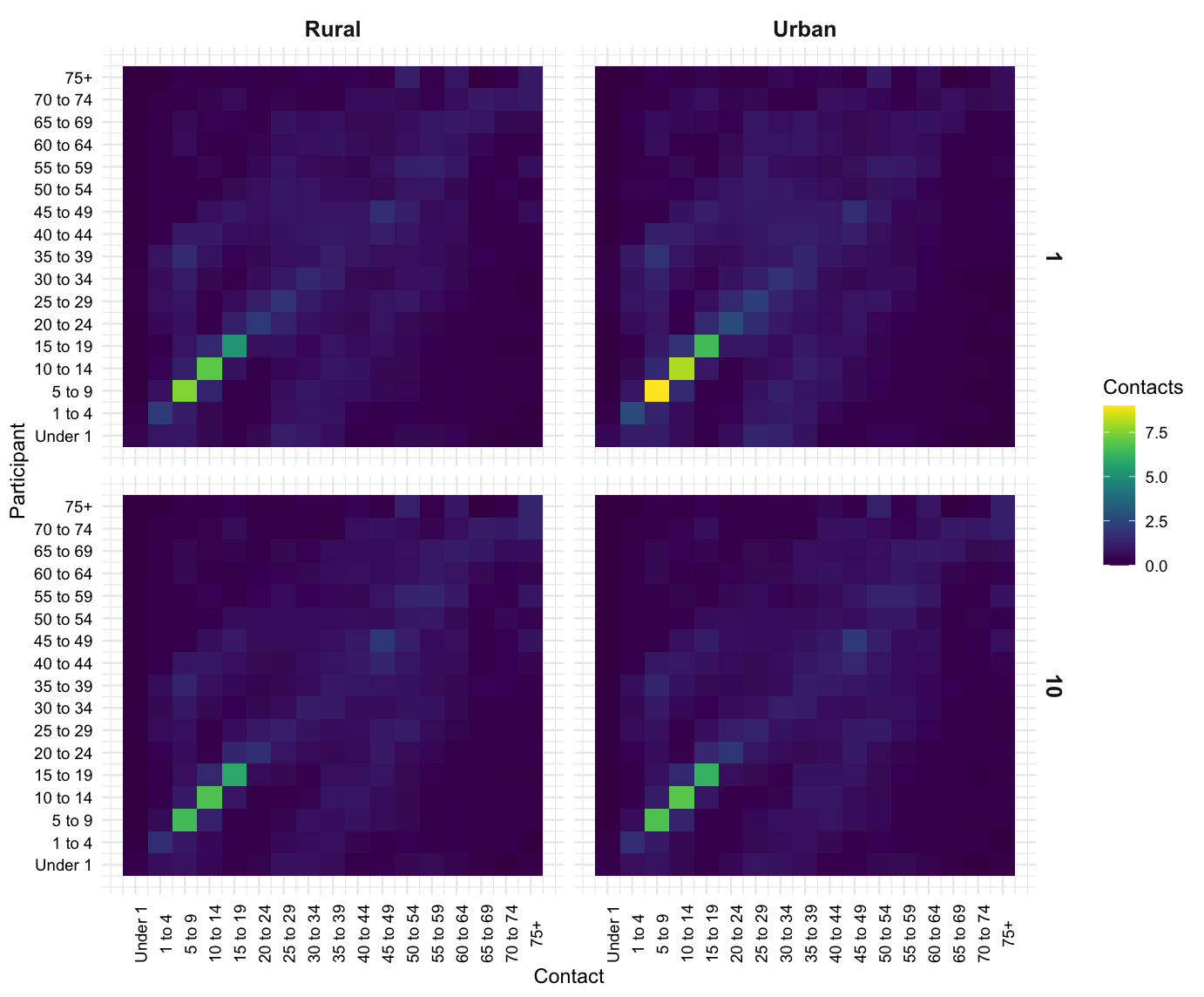

**Supplementary Figure 2.** Projected contact matrices for urban and rural areas of the IMD deciles 1 and 10.

### 3. Age- and IMD-specific health statuses

Supplementary Figure 3 shows the proportion of IMD deciles 1 and 10 self-reporting as in ‘Very good’, ‘Good’, ‘Fair’, ‘Bad’, or ‘Very bad’ health in each age group. Those living in the most deprived 10% of areas visibly spend a greater time in ill-health than those in the least deprived 10%. Health prevalence was then calculated as the proportion of the population in the upper two sections of the graph (‘Very good’, ‘Good’ health).

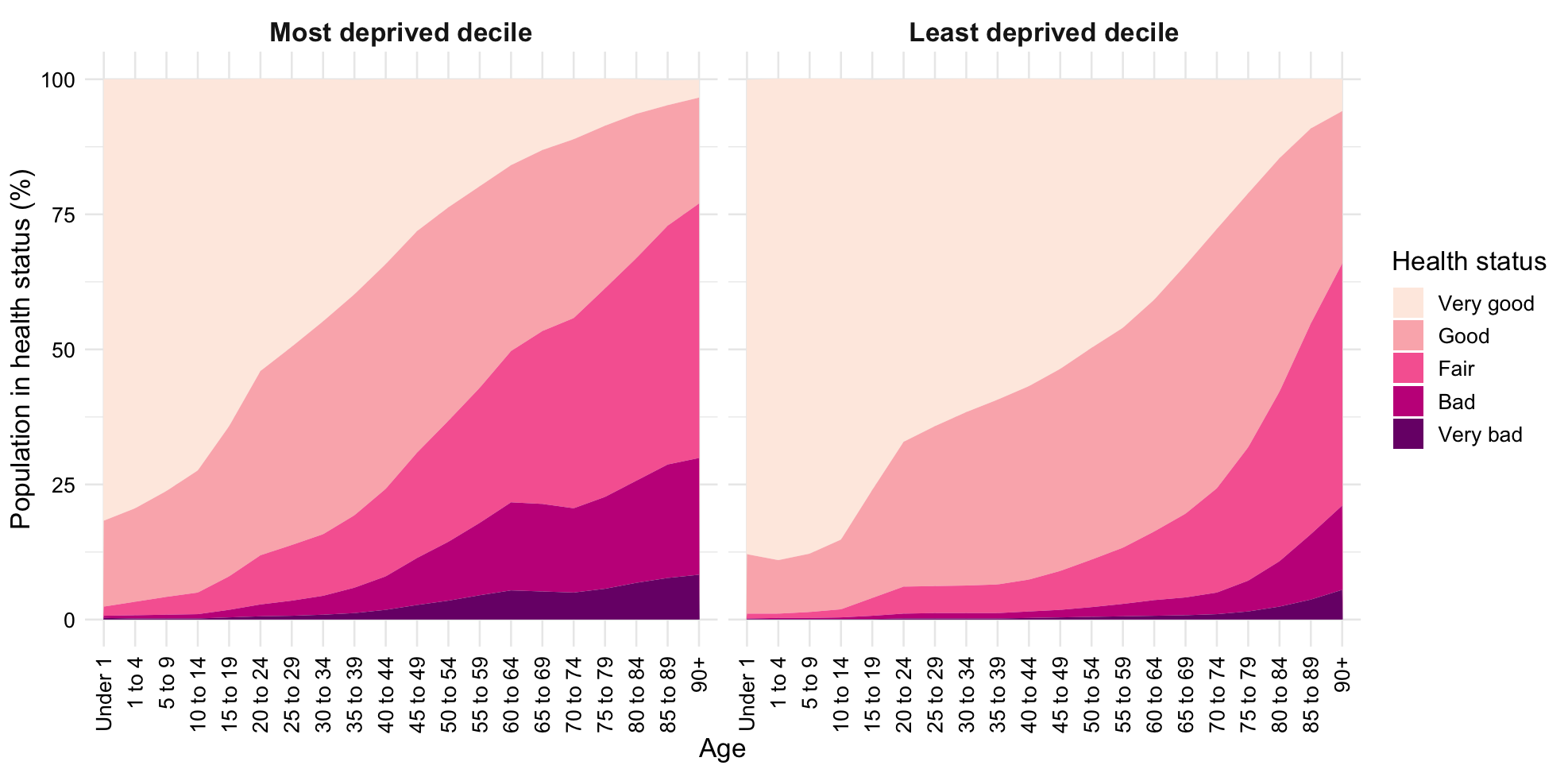

**Supplementary Figure 3.** Age-specific health statuses in the most and least deprived deciles in England. Data from Census 2021.

### 4.  Overall health prevalence in each IMD decile

Supplementary Table 2 shows the overall proportion of each IMD decile self-reporting as in ‘Very good’ or ‘Good’ health (overall health prevalence). There is a consistent decrease in health prevalence as relative deprivation increases, and health prevalence is 11% higher in the least deprived decile than the most deprived. Despite more deprived areas having younger populations on average (see Figure 1a, Supplementary Figure 1), the overall health prevalence is still lower.

| **IMD decile** | 1 (Most deprived) | 2 | 3 | 4 | 5 | 6 | 7 | 8 | 9 | 10 (Least deprived) |
| --- | --- | --- | --- | --- | --- | --- | --- | --- | --- | --- |
| **Health prevalence** | 0.76 | 0.79 | 0.80 | 0.81 | 0.82 | 0.83 | 0.84 | 0.85 | 0.85 | 0.87 |

**Supplementary Table 2.** Overall health prevalence in each IMD decile.

### 5. Training dataset

Supplementary Table 3 shows the age-specific health prevalence and clinical fraction estimates for the general population of England, in ten-year age groups, used as a training dataset for the function between the two. In the training dataset, older age groups have consistently lower health prevalences and higher clinical fractions.

| **Age group** | **10-19** | **20-29** | **30-39** | **40-49** | **50-59** | **60-69** | **70+** |
| --- | --- | --- | --- | --- | --- | --- | --- |
| **Health prevalence** | 0.95 | 0.91 | 0.89 | 0.84 | 0.77 | 0.70 | 0.54 |
| **Clinical fraction** | 0.21 | 0.27 | 0.33 | 0.40 | 0.49 | 0.63 | 0.69 |

**Supplementary Table 3.** Age-specific health prevalence and clinical fraction estimates for the general population of England, in ten-year age groups, used as a training dataset for the function between the two.

### 6. Clinical mortality probabilities

The age-specific likelihood of an individual assigned to the clinical arm of the model being assigned the D compartment (as opposed to R),
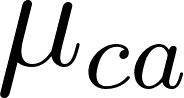
<https://www.codecogs.com/eqnedit.php?latex=%5Cmu_%7Bca%7D#0>, was calculated by dividing the IFR
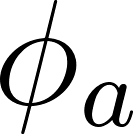
<https://www.codecogs.com/eqnedit.php?latex=%5Cphi_a#0> by the clinical fraction
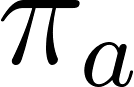
<https://www.codecogs.com/eqnedit.php?latex=%5Cpi_a#0>. Supplementary Table 4 shows the resulting age-specific clinical mortality probabilities, which range from 0.00555% in those younger than 10 years old to 11.3% in those aged 80+. For the age group 75+ in the transmission model, the mean of the 70-79 and 80+ age groups was used (0.0875).

| **Age** | **0-9** | **10-19** | **20-29** | **30-39** | **40-49** | **50-59** | **60-69** | **70-79** | **80+** |
| --- | --- | --- | --- | --- | --- | --- | --- | --- | --- |
| 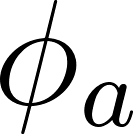<https://www.codecogs.com/eqnedit.php?latex=%5Cphi_a#0> | 0.0000161 | 0.0000695 | 0.000309 | 0.000844 | 0.00161 | 0.00595 | 0.0193 | 0.0428 | 0.0780 |
| 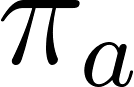<https://www.codecogs.com/eqnedit.php?latex=%5Cpi_a#0> | 0.29 | 0.21 | 0.27 | 0.33 | 0.40 | 0.49 | 0.63 | 0.69 | 0.69 |
| 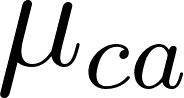<https://www.codecogs.com/eqnedit.php?latex=%5Cmu_%7Bca%7D#0> | 0.0000555 | 0.000331 | 0.00114 | 0.00256 | 0.00403 | 0.0121 | 0.0306 | 0.0620 | 0.113 |

**Supplementary Table 4.** Age-specific infection fatality ratios from [[1]](https://www.zotero.org/google-docs/?eZ4aIV), clinical fractions from [[2]](https://www.zotero.org/google-docs/?uSWqtd), and corresponding clinical mortality probabilities.

### 7.  Model equations

The epidemic model as shown in Figure 1c is described by the below equations, which detail age-specific movement between the model compartments in a given IMD- and geography-specific population.

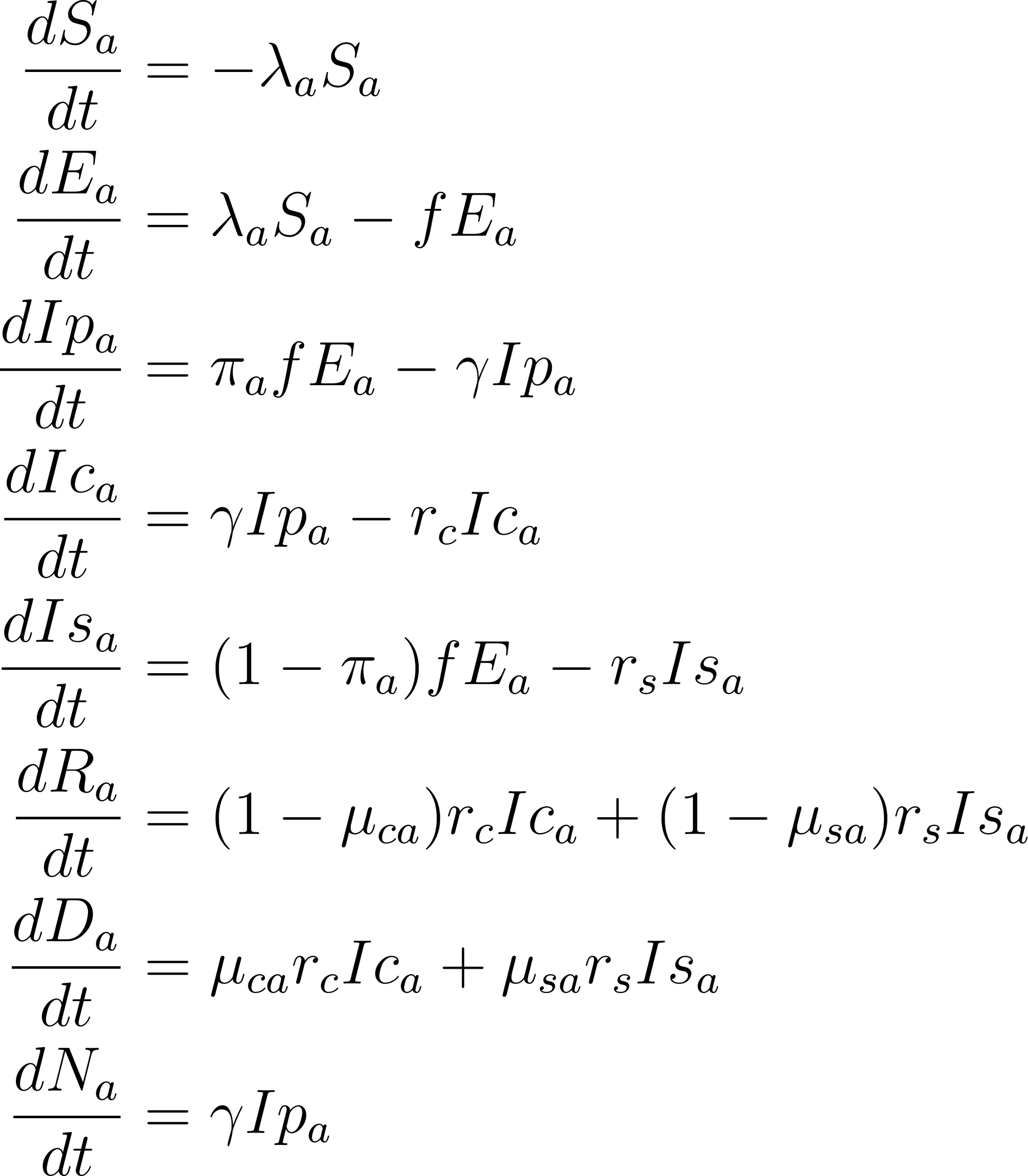
<https://www.codecogs.com/eqnedit.php?latex=%20%5Cbegin%7Baligned%7D%20%5Cfrac%7BdS_a%7D%7Bdt%7D%20%26%3D%20-%20%5Clambda_a%20S_a%20%5C%5C%5C%5C%20%5Cfrac%7BdE_a%7D%7Bdt%7D%20%26%3D%20%5Clambda_a%20S_a%20-%20f%20E_a%20%5C%5C%5C%5C%20%5Cfrac%7BdIp_a%7D%7Bdt%7D%20%26%3D%20%5Cpi_a%20f%20E_a%20-%20%5Cgamma%20Ip_a%20%5C%5C%5C%5C%20%5Cfrac%7BdIc_a%7D%7Bdt%7D%20%26%3D%20%5Cgamma%20Ip_a%20-%20r_c%20Ic_a%20%5C%5C%5C%5C%20%5Cfrac%7BdIs_a%7D%7Bdt%7D%20%26%3D%20(1%20-%20%5Cpi_a)%20f%20E_a%20-%20r_s%20Is_a%20%5C%5C%5C%5C%20%5Cfrac%7BdR_a%7D%7Bdt%7D%20%26%3D%20(1-%5Cmu_%7Bca%7D)r_c%20Ic_a%20%2B%20(1%20-%20%5Cmu_%7Bsa%7D)%20r_s%20Is_a%20%5C%5C%5C%5C%20%5Cfrac%7BdD_a%7D%7Bdt%7D%20%26%3D%20%5Cmu_%7Bca%7D%20r_c%20Ic_a%20%2B%20%5Cmu_%7Bsa%7D%20r_s%20Is_a%20%5C%5C%5C%5C%20%5Cfrac%7BdN_a%7D%7Bdt%7D%20%26%3D%20%5Cgamma%20Ip_a%20%5Cend%7Baligned%7D#0>

As the model has a total population of 1, the size of each compartment represents the proportion of the population in that compartment. In the initial state of the epidemic, all individuals are susceptible except for 0.1% of the population assigned to the Ip compartment in age group 30 - 34, to represent an infection brought in by an individual likely travelling for work. The susceptible initial conditions are therefore the age distribution
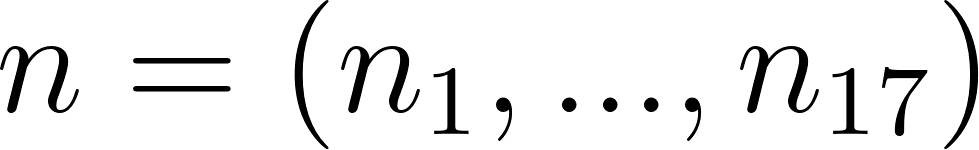
<https://www.codecogs.com/eqnedit.php?latex=n%20%3D%20(n_1%2C...%2Cn_%7B17%7D)#0>, with a reduction of 0.001 in the age group 30 - 34 (
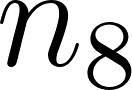
<https://www.codecogs.com/eqnedit.php?latex=n_8#0>). The model also has a compartment counting age-specific cumulative clinical cases,
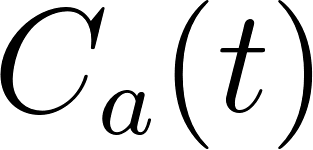
<https://www.codecogs.com/eqnedit.php?latex=C_a(t)#0>, which does not affect any transmission dynamics and has initial conditions
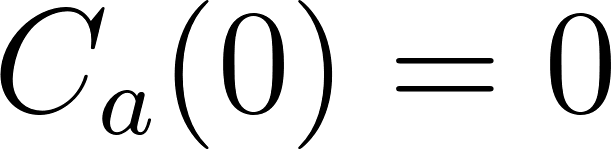
<https://www.codecogs.com/eqnedit.php?latex=C_a(0)%20%3D%200#0> for all a.

### 8. Epidemic trajectories

Supplementary Figure 4a shows the total infections on each day of the epidemic (*Ip* + *Ic* + *Is*, aggregated over the 17 age groups) for each IMD decile in both geographies, and indicates a larger peak of infections occurring on an earlier day of the epidemic in more deprived areas, in both urban and rural areas. The cumulative clinical cases of epidemics in each IMD decile and geography show a greater number of clinical cases and earlier epidemics in more deprived areas (Supplementary Figure 4b). Rural areas experience lower peaks of infections than urban areas in this model, but more clinical cases.

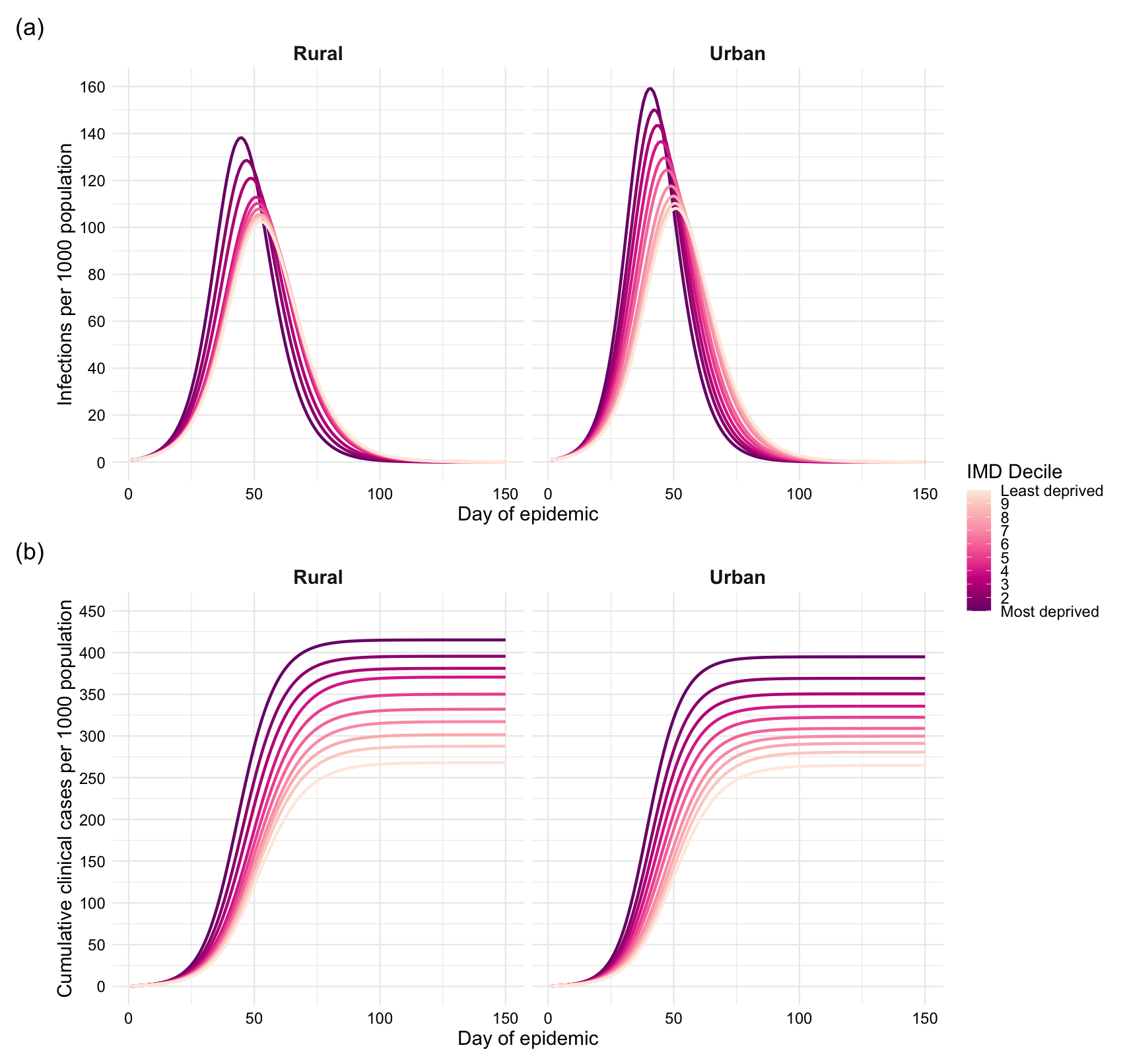

**Supplementary Figure 4.** **a)** Infections and **b)** cumulative clinical cases, per 1,000 population, over the first 150 days of a COVID-19 epidemic in each IMD decile, stratified by geography.

### 9. Counterfactual scenario of constant age structure

Using an age structure independent of IMD decile or geography, each of the measures of epidemiological burden is consistently higher for more deprived areas (Supplementary Figure 5). The model predicts 52 more infections, 156 more clinical cases, and 2.4 more deaths per 1,000 population in the most than least deprived decile (Supplementary Figure 5a, b, d), an IFR 0.24% higher (Supplementary Figure 5e), 18.9 more clinical cases per 1,000 population at the clinical peak of the epidemic (Supplementary Figure 5c), and 0.21 increase in R0 (Supplementary Figure 5f).

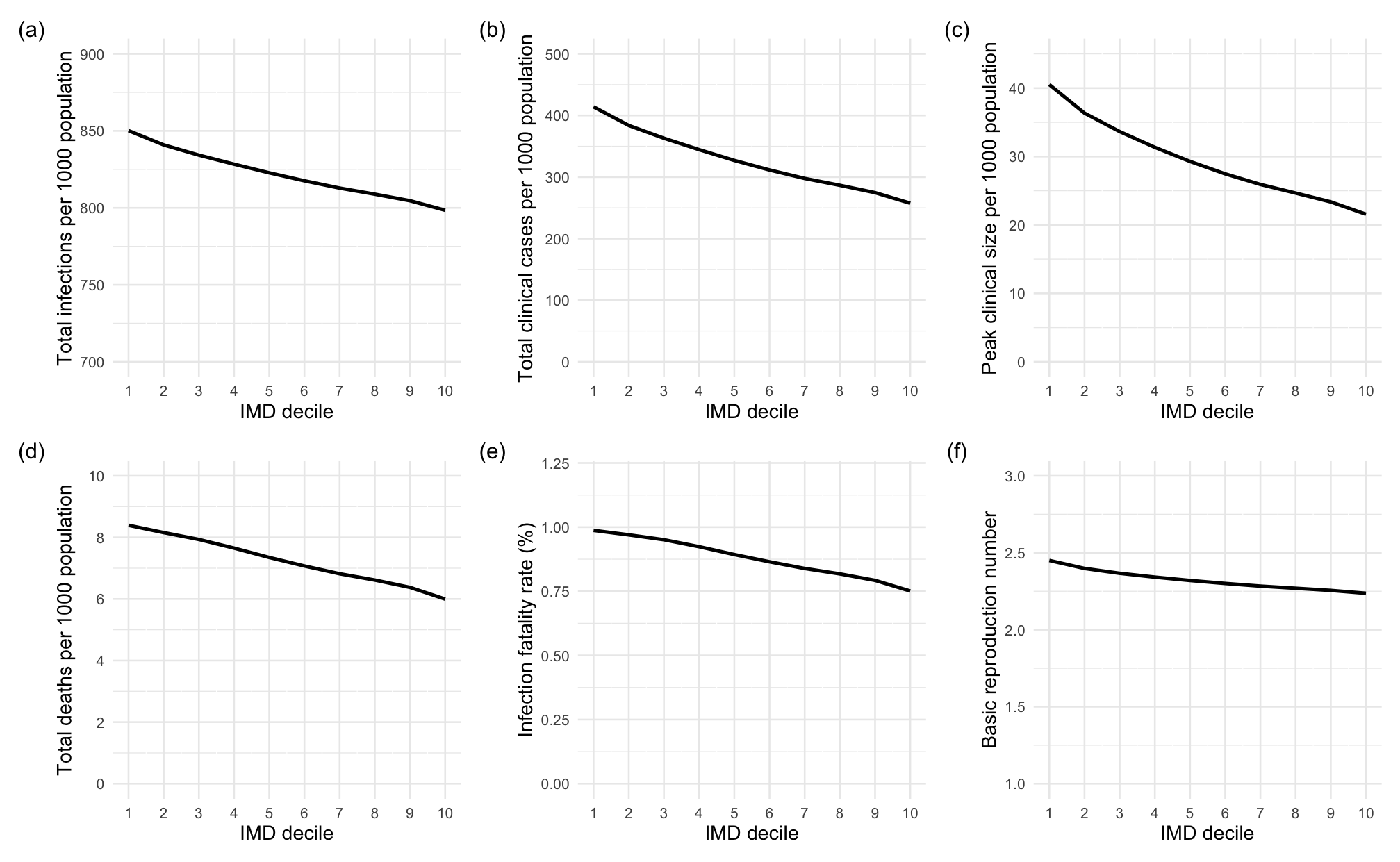

**Supplementary Figure 5.** Measures of size of a COVID-19 epidemic in each IMD decile, assuming a constant age structure, where 1 is the most deprived decile and 10 the least deprived. **a)** Total infections, **b)** total clinical cases, **c)** peak clinical size, **d)** total deaths, all per 1,000 population, **e)** infection fatality ratio, **f)** basic reproduction number.

### 10. Counterfactual scenario of underlying health equity

Under the assumption that all IMD deciles experience the same age-specific clinical fractions as the least deprived decile, deaths are prevented at all ages. Supplementary Figure 6 shows the scaled-up number of deaths attributable to underlying health inequity in each age group (i.e., those which occur under the original model assumptions, but not in the counterfactual scenario of underlying health equity). Supplementary Figure 7 shows the percentage of deaths which occurred under the original model conditions which are attributable to underlying health. Improved underlying health is most effective at reducing mortality between ages 30 and 70; almost 35% of deaths occurring in the age group 50-54 are prevented by underlying health equity. The decreased effectiveness of health equity in the oldest age groups is likely due to the convergence in health prevalence in those aged 70+, which is over-exaggerated due to the upper bound set on the clinical fraction in this model which are reached in some IMD deciles in ages 60+. Therefore, this model likely underestimates the effectiveness of underlying health equity in those aged 60+, and will therefore likely underestimate the proportion of overall deaths which would be prevented by improved health.

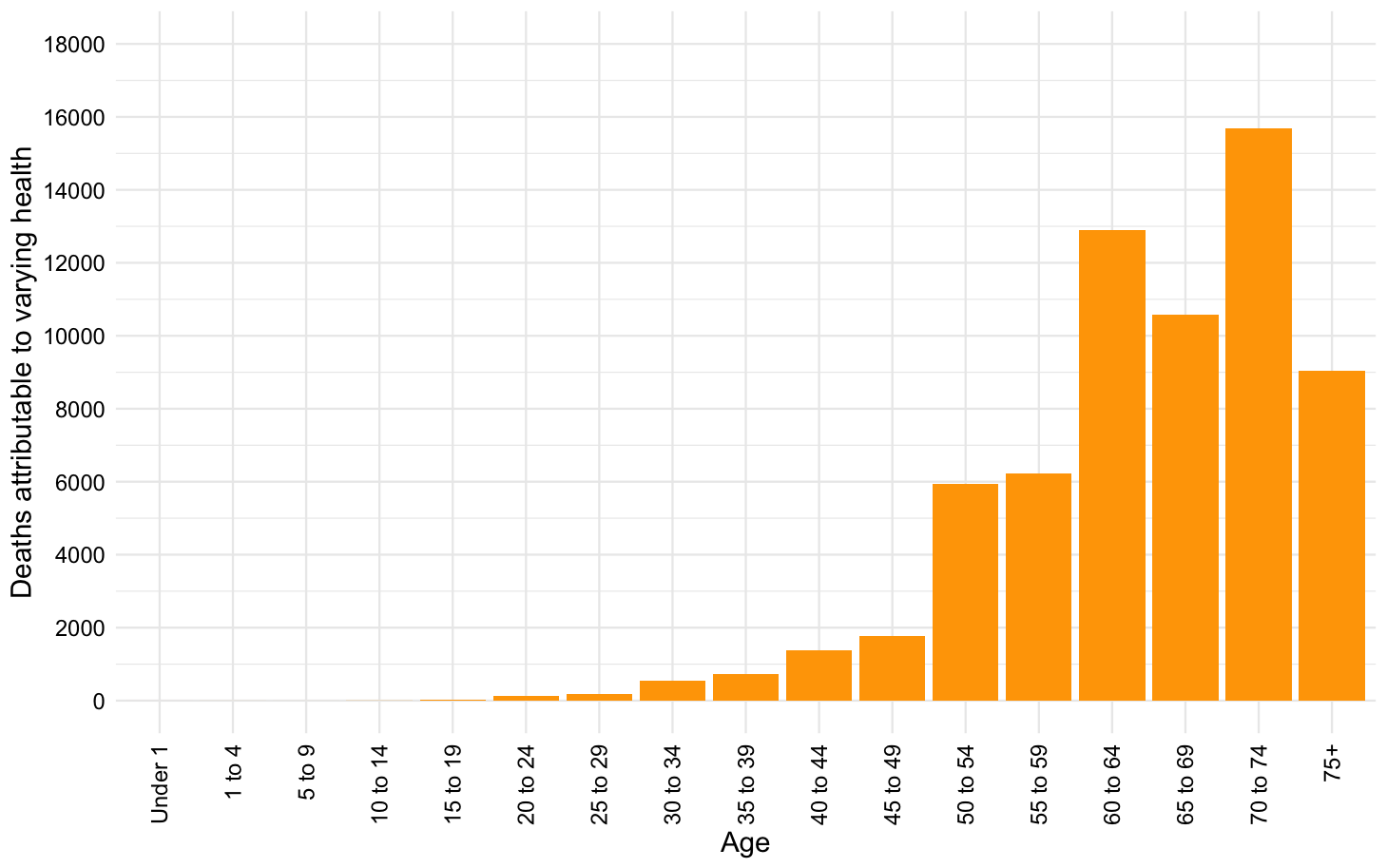

**Supplementary Figure 6.** Deaths occurring in each age group, scaled up across England, which are prevented in the case of underlying health equity.

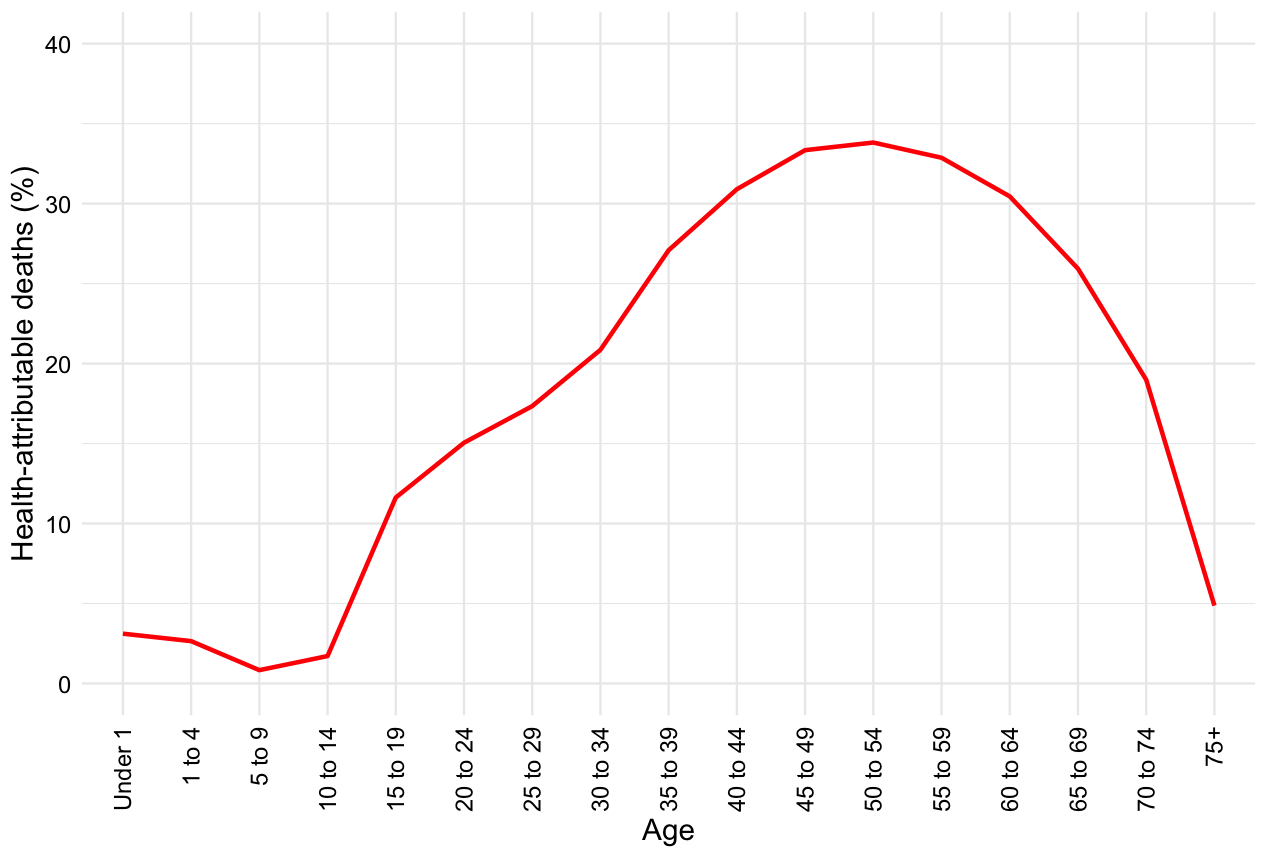

**Supplementary Figure 7.** Age-specific percentage of deaths occurring under the original model conditions, scaled up across England, which are prevented in the case of underlying health equity.

### 11.  School closures

To examine the effectiveness of school closures in reducing mortality under different conditions, we varied the threshold for school closure implementation. Initially, school closures were implemented once 5% of the population had developed a clinical case of COVID-19 (P = 0.05). Supplementary Figure 8a shows the reduction in deaths in urban and rural areas in each IMD decile as P ranges from 0 to 0.3. The most school closures with the greatest mortality reductions are those implemented earlier, ideally when P = 0. As P increases and closures are implemented later in the epidemic, more deprived areas begin to experience greater mortality reductions than affluent areas, reversing the inequality seen in Figure 5c. This is likely because more affluent areas experience smaller epidemics overall, and so thresholds such as 25% of the population experiencing a clinical case of COVID-19 are more effective in areas where the epidemic is still developing than those where the epidemic has already peaked.

In order to control for this effect, we also used a time-based threshold which triggered school closures a fixed number of days after the first case was introduced, ranging from day 10 to day 50. The pattern of inequality now remains unchanged by different times of implementation, and only the magnitude of effectiveness varies (Supplementary Figure 8b). The most effective intervention is the still earliest possible one.

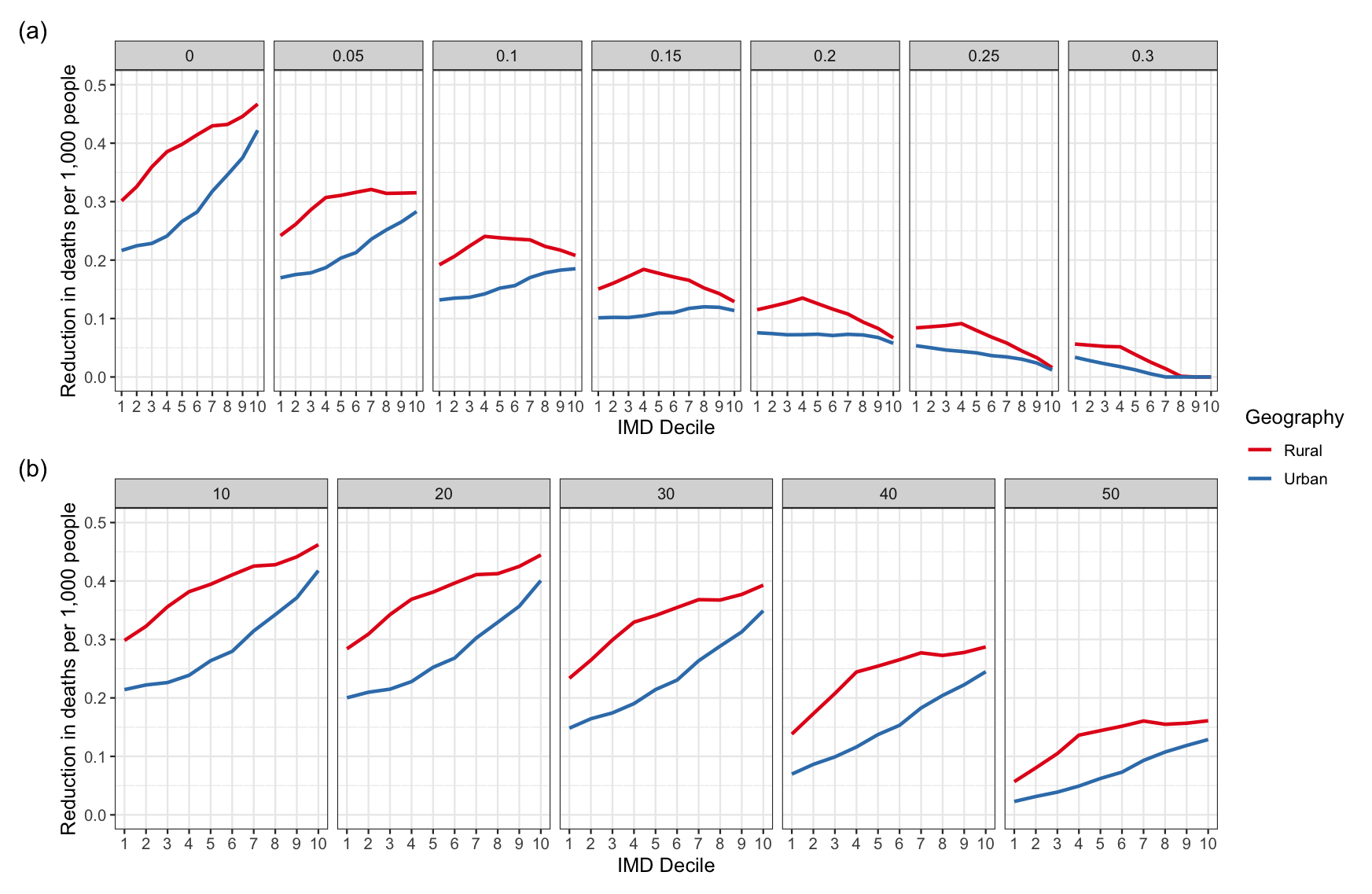

**Supplementary Figure 8. Mortality under varying school closure implementation thresholds. a)** IMD- and geography-specific reductions in the crude number of deaths observed per 1,000 population, split by the implementation threshold *P*. **b)** IMD- and geography-specific reductions in the crude number of deaths observed per 1,000 population, split by the time-based implementation threshold in days.

We also explored the reductions in *R_0_* when holding either underlying health or age constant within a given geography (Supplementary Figure 9). When clinical fraction was constant, very similar reductions in *R_0_* occurred to in the original model. When age structure was held constant across all IMD deciles, school closures resulted in greater reductions in *R_0_* in more affluent areas, showing the effect of clinical fraction alone. The original U-shape in *R_0_* reductions is likely to be mainly caused by the age structure of each population, with a slight effect of clinical fraction too.

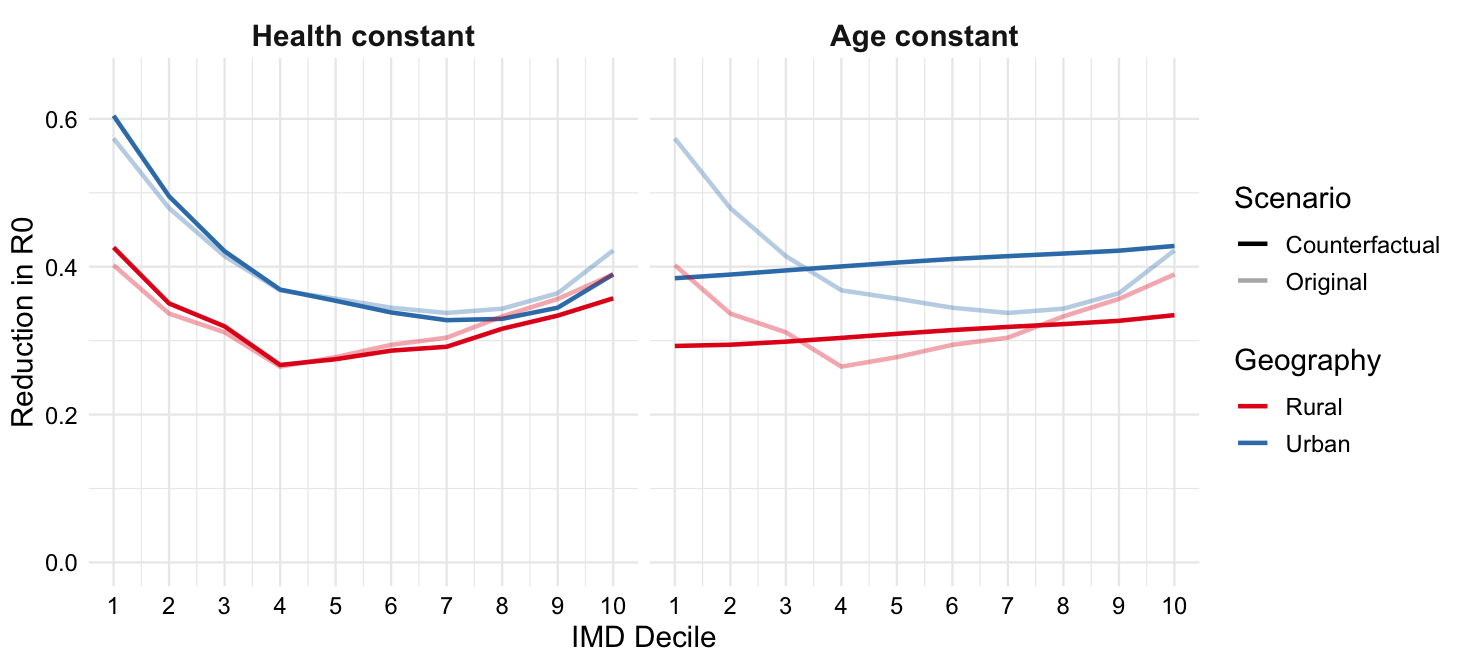

**Supplementary Figure 9.** The reduction in *R_0_* after school closures, with health prevalence held constant, or age structure held constant (reduction in *R_0_* under the original model assumptions shown as pale lines).

### 12. Epidemiological sensitivity analyses

We analysed mortality rates when varying the dependence of clinical fraction on health prevalence. For this analysis, age-specific clinical fractions were calculated by

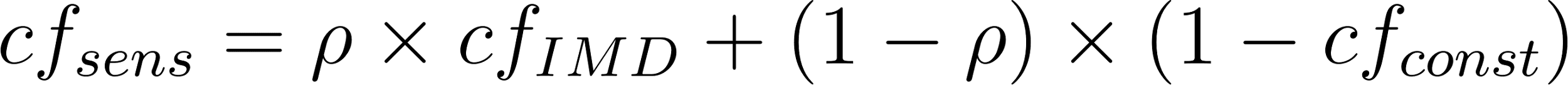
<https://www.codecogs.com/eqnedit.php?latex=cf_%7Bsens%7D%20%3D%20%5Crho%20%20%5Ctimes%20cf_%7BIMD%7D%20%2B%20(1-%5Crho)%20%5Ctimes%20(1-cf_%7Bconst%7D)#0>,

for
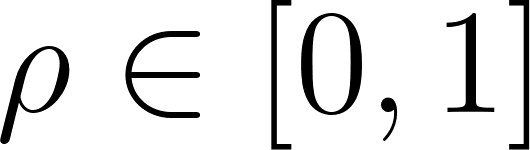
<https://www.codecogs.com/eqnedit.php?latex=%5Crho%20%5Cin%20%5B0%2C1%5D#0>, where
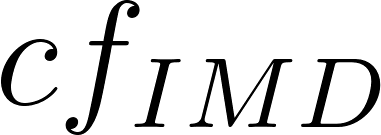
<https://www.codecogs.com/eqnedit.php?latex=cf_%7BIMD%7D#0> is the IMD-specific clinical fraction used previously and
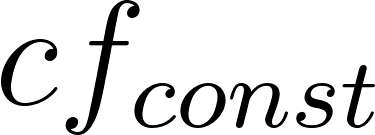
<https://www.codecogs.com/eqnedit.php?latex=cf_%7Bconst%7D#0> is the clinical fraction found by Davies et al. [[2]](https://www.zotero.org/google-docs/?cWKDsJ) and is hence independent of IMD. Supplementary Figure 10 shows how the distribution of deaths (crude and age-standardised by geography) across IMD deciles changes with dependence. As the age-specific clinical fractions become less dependent on health prevalence (
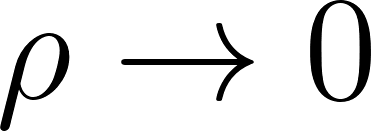
<https://www.codecogs.com/eqnedit.php?latex=%5Crho%20%5Cto%200#0>), fewer deaths (crude and age-standardised) occur in the most deprived 40% of areas, and more occur in the most affluent 60% of areas, in both urban and rural geographies.

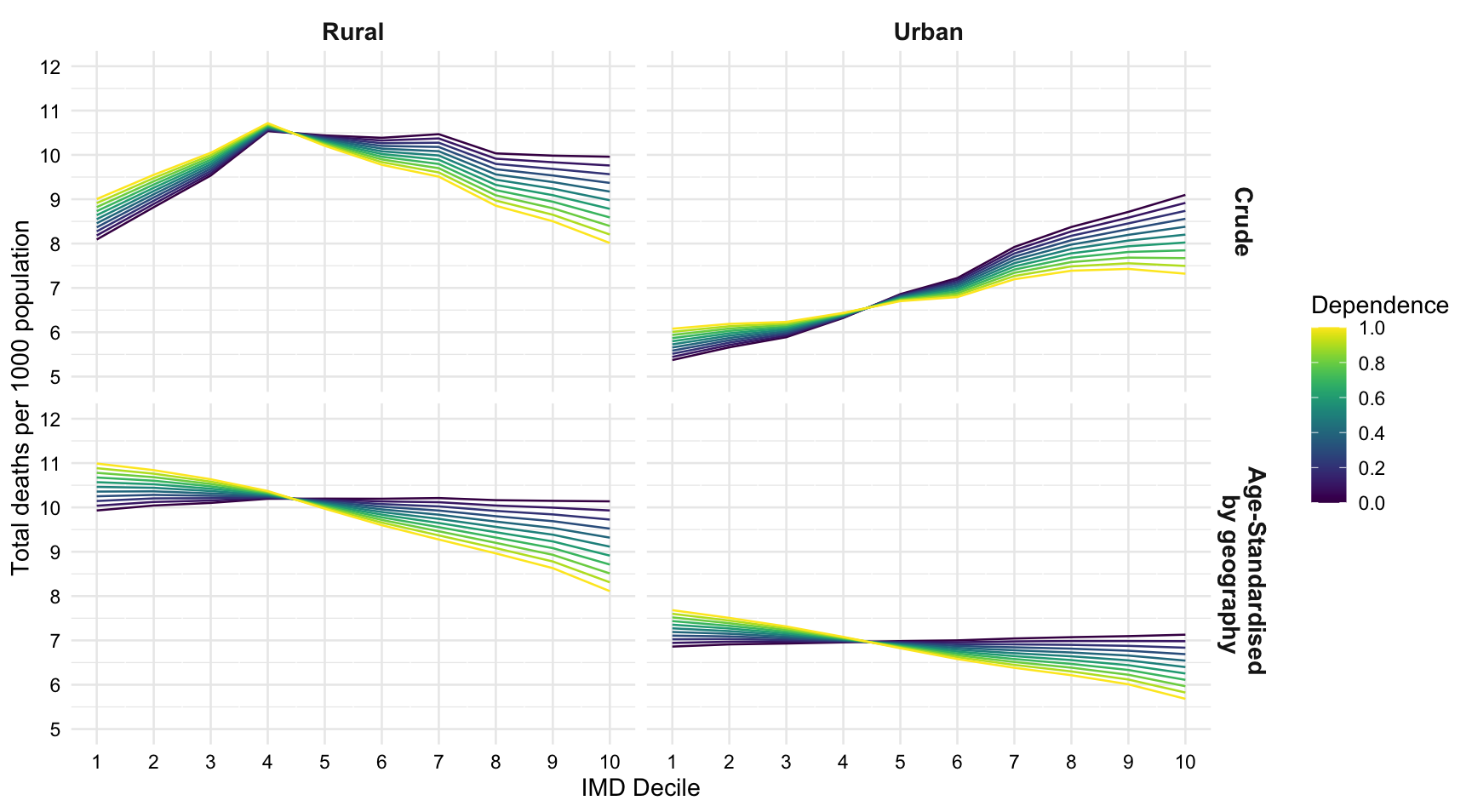

**Supplementary Figure 10.** IMD- and geography-specific deaths per 1,000 population, crude and age-standardised by geography, as the dependence of clinical fraction on underlying health (
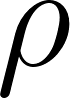
<https://www.codecogs.com/eqnedit.php?latex=%5Crho#0>) varies.

While the relative subclinical infectiousness was assumed to be
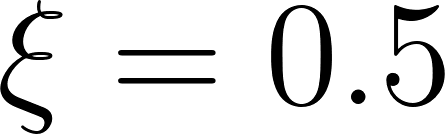
<https://www.codecogs.com/eqnedit.php?latex=%5Cxi%20%3D%200.5#0> in the initial model, we conducted a sensitivity analysis to determine how varying
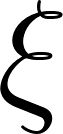
<https://www.codecogs.com/eqnedit.php?latex=%5Cxi#0> between 0 and 1 affects the epidemiological outcomes. The distributions of total infections and total clinical cases across deprivation deciles and geography were largely unaffected by the value of
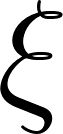
<https://www.codecogs.com/eqnedit.php?latex=%5Cxi#0>, and only changed in magnitude: lower
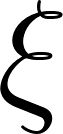
<https://www.codecogs.com/eqnedit.php?latex=%5Cxi#0> corresponded to fewer total infections and clinical cases. Supplementary Figure 11 shows deaths (crude and age-standardised by geography) per 1,000 population. Age-standardised mortality follows a similar distribution for each value of
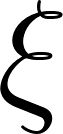
<https://www.codecogs.com/eqnedit.php?latex=%5Cxi#0>, with a steeper gradient of inequality in mortality by deprivation as
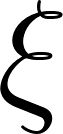
<https://www.codecogs.com/eqnedit.php?latex=%5Cxi#0> decreases. The crude number of deaths is more affected by age distribution for greater
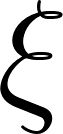
<https://www.codecogs.com/eqnedit.php?latex=%5Cxi#0>, but more affected by clinical vulnerability for low
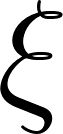
<https://www.codecogs.com/eqnedit.php?latex=%5Cxi#0>. This means that if subclinical cases are roughly as infectious as clinical cases, more crude deaths are observed in more affluent areas, but in the case that subclinical cases are relatively much less infectious then more crude deaths are observed in more deprived areas.

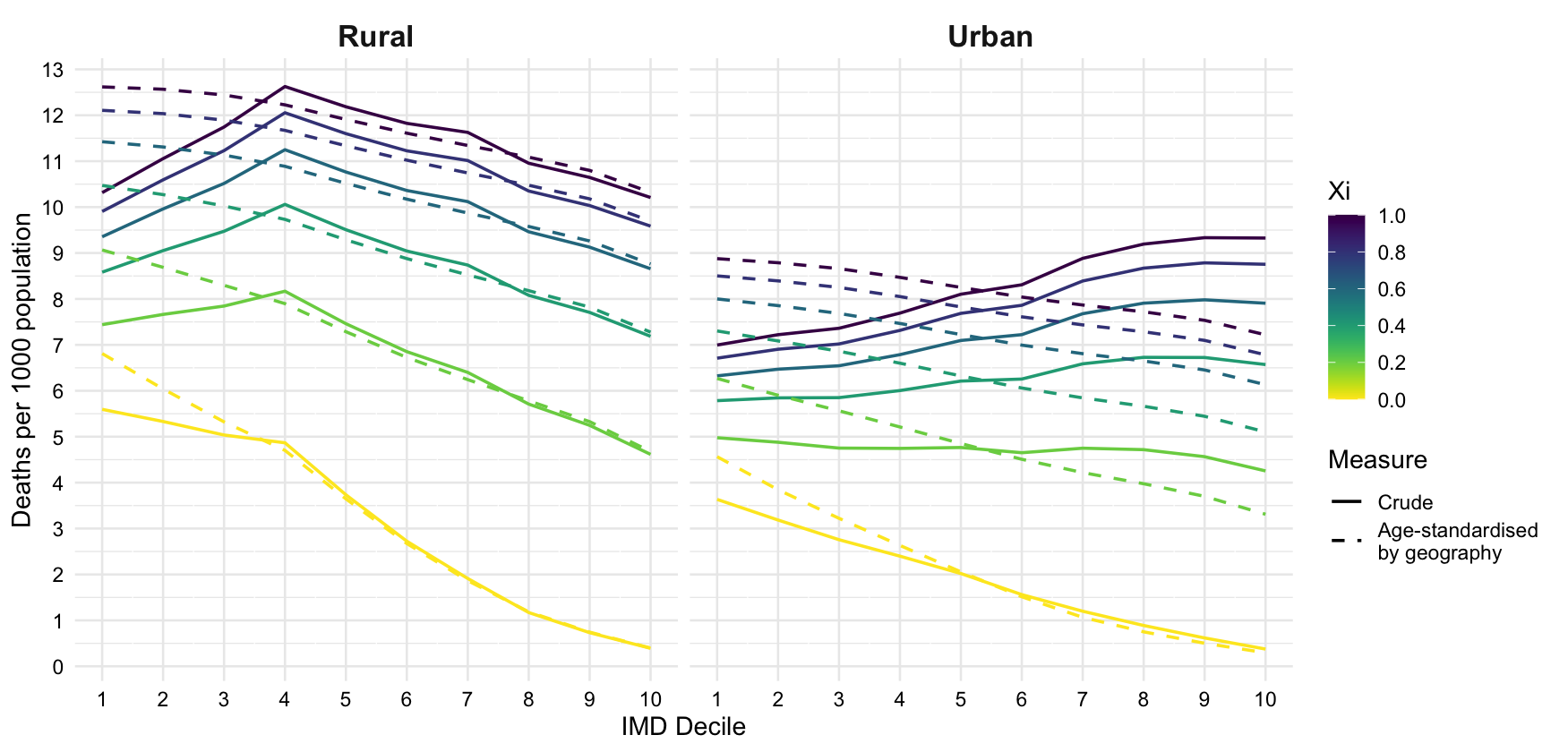

**Supplementary Figure 11.** IMD- and geography-specific deaths per 1,000 population, crude and age-standardised by geography, as relative subclinical infectiousness varies.

An analysis of the impact of varying the length of the epidemiological periods showed very similar results to that of varying relative subclinical infectiousness. By multiplying each epidemiological period by a scalar *a* in [0.4, 3], and therefore changing the parameters to

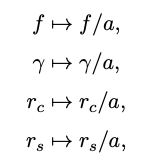

much shorter or longer epidemics with different basic reproduction numbers were investigated. In the case of a = 0.4, the infectious period is reduced by 60%, reducing R0 and meaning that only the most deprived 20% of urban areas experience epidemics with R0 > 1. In the case of a = 3, infectious periods are tripled and R0 now ranges between 6.3 and 8.1. In terms of inequalities, total infections, total clinical cases, and age-standardised deaths change magnitude but not distribution, similar to above. Crude mortality’s dependence on age structure is related to the length of the epidemiological periods (Supplementary Figure 12). In the case of increased infection periods (a > 1), the crude number of deaths closely relates to median age, but as epidemiological periods shorten (a < 1), crude mortality decreases monotonously with affluence in both geographies and reflects clinical vulnerability.

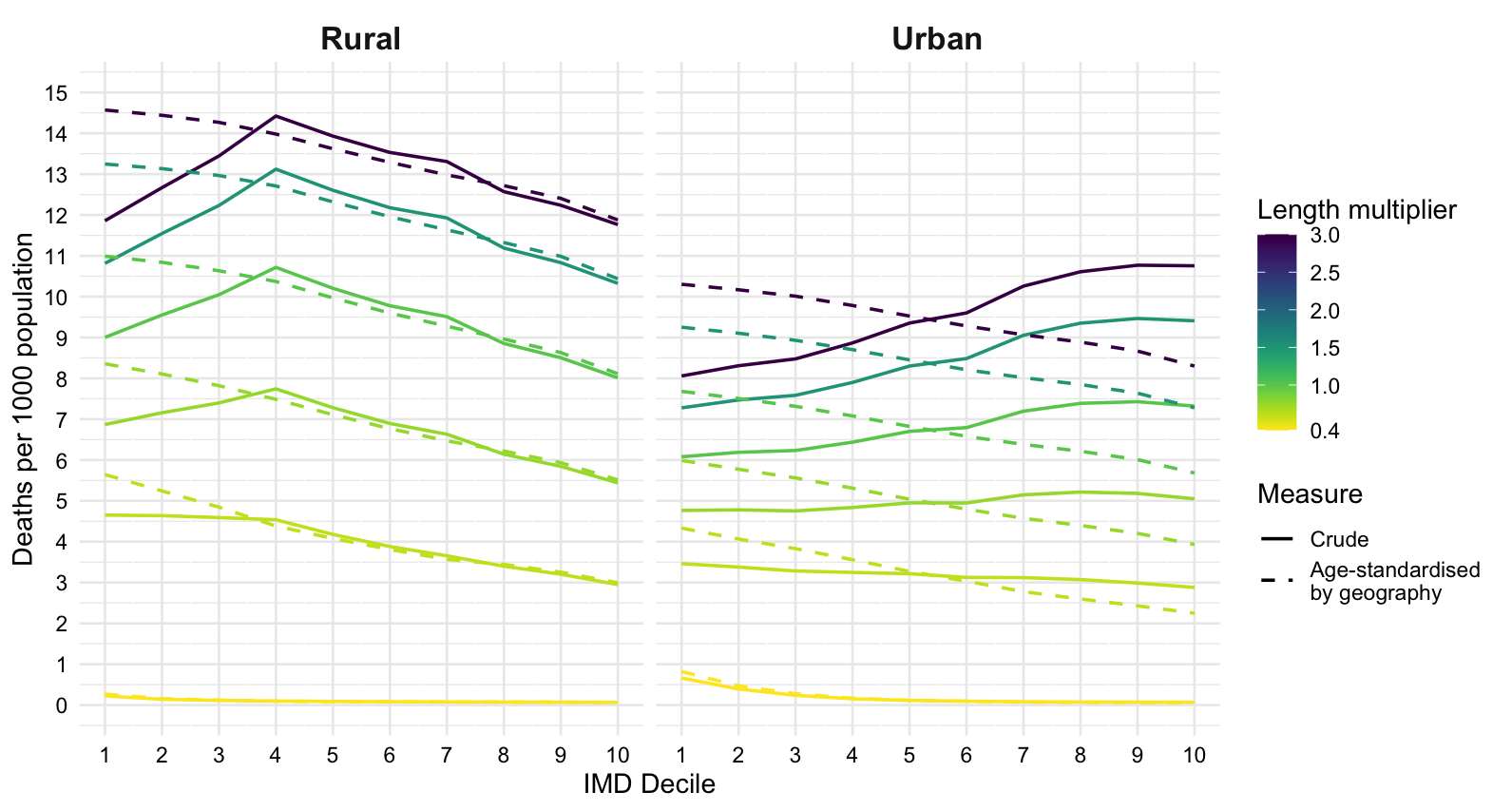

**Supplementary Figure 12.** IMD- and geography-specific deaths per 1,000 population, crude and age-standardised by geography, as the length of the epidemiological periods varies.

### 13. Association between health prevalence and IMD domains at LSOA level

We investigated the relationships between health prevalence and the seven domains of the IMD by comparing the crude health prevalence in each LSOA to its rankings in each domain of deprivation. Rankings (and deciles) for each LSOA were published by the Ministry of Housing, Communities & Local Government in 2019 [[3]](https://www.zotero.org/google-docs/?tmnzem). The dataset of health states for each LSOA (without age-stratification) was downloaded from the ONS website and converted into overall health prevalence [[4]](https://www.zotero.org/google-docs/?bY5zDB). The two datasets were linked via LSOA codes, allowing for comparisons between rankings in each domain of deprivation (ranging from 1, most deprived, to 32,844, least deprived) and health prevalence.

Health prevalence is associated with IMD, but also its components; Supplementary Figure 13 shows the association of health prevalence with each IMD domain, with overall IMD rank determining the colour of each data-point. High ranks represent the least deprived LSOAs. The strongest association with health prevalence is seen between rankings in income, employment, education, and health and disability. There are less strong correlations between crime, barriers to housing and services, and living environment and health prevalence. While the rankings are relative and not absolute measures of the domains (as with IMD deciles themselves), and the health prevalences are affected by underlying age structure, this approach is insightful for confirming that domains of deprivation other than health and disability are associated with health prevalence.

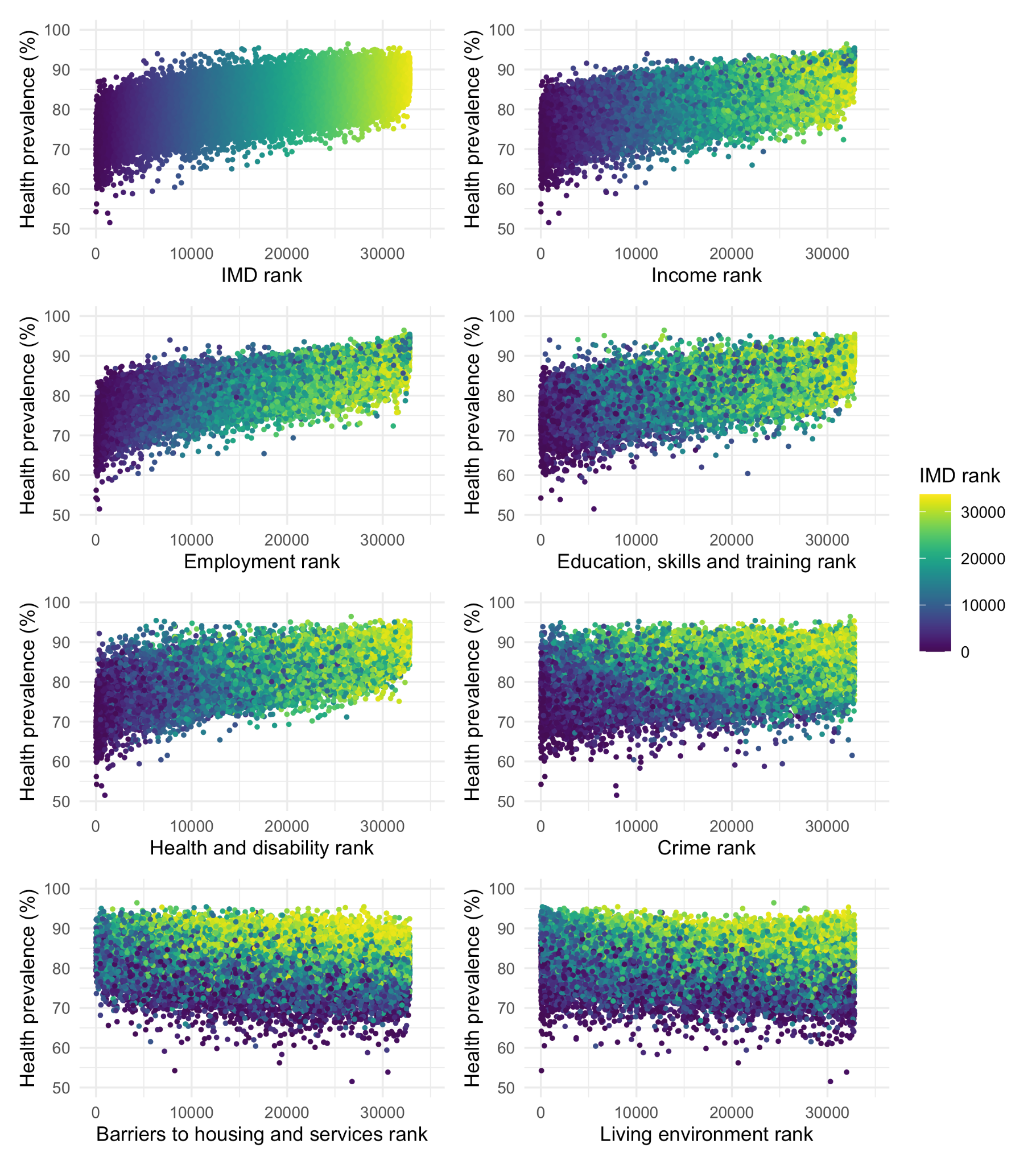

**Supplementary Figure 13.** Relationship between the ranks of LSOAs in each IMD domain (1 the most deprived, 32,844 the least deprived) and health prevalence.
